# Migration-linked tuberculosis epidemics: how the future of tuberculosis in high-income countries will depend on the success or failure of TB control in high-burden settings

**DOI:** 10.1101/2025.11.14.25340268

**Authors:** Byron LM Cohen, Kevin Croke, William P Hanage, Ted Cohen, Nicolas A Menzies

**Affiliations:** Department of Global Health and Population, Harvard TH Chan School of Public Health, Boston MA, USA; Horizon Institute for Public Service, Washington, DC, USA; Department of Epidemiology, Harvard TH Chan School of Public Health, Boston MA, USA; Department of Epidemiology of Microbial Diseases, Yale School of Public Health, New Haven, CT, USA; Center for Health Decision Science, Harvard TH Chan School of Public Health, Boston MA, USA

## Abstract

**Background:** Globally, *Mycobacterium tuberculosis* (*Mtb*) is the leading cause of death due to a single pathogen, with most tuberculosis cases occurring in low- and middle-income countries. A growing proportion of tuberculosis cases in high-income countries occur among foreign-born individuals, often resulting from a *Mtb* infection acquired before migration. As a result, tuberculosis trends in many high-income countries are increasingly influenced by tuberculosis epidemiology in and migration patterns from other countries. Our objective was to estimate how the future risks of tuberculosis in high-income countries will change depending on the success or failure of efforts to combat tuberculosis in high-burden settings.

**Methods:** We defined scenarios representing different levels of optimism regarding tuberculosis control in high-burden settings. The most optimistic scenario assumed high-burden countries would achieve tuberculosis elimination targets proposed by the WHO. The most pessimistic assumed major increases in tuberculosis following sharp reductions in international health aid. A base-case scenario assumed continuation of pre-2025 trends. We used calibrated mathematical models to predict how these scenarios would change *Mtb* infection prevalence among future migrants and thereby affect tuberculosis incidence and deaths in high-income countries. We considered 49 high-burden countries (as defined by the WHO), and projected tuberculosis outcomes in 60 high-income countries until 2050.

**Findings:** Over 2025-2050, we project there will be 2,266,000 (95% credible interval (CI): 1,938,000–2,744,000) tuberculosis cases in high-income countries if pre-2025 trends continue, with 57% (95%CI: 50–65) of these cases occurring among foreign-born individuals (up from 39% in 2024), for an average incidence rate of 6.5 (95%CI: 5.3–8.3) per 100,000 in 2050. Under the most optimistic scenario we estimated that there would be 785,000 (95%CI: 647,000–950,000) fewer tuberculosis cases and 63,000 (95%CI: 52,000–78,000) fewer tuberculosis deaths in high-income countries over 2025-2050, with an incidence of 2.3 (95%CI: 1.8–3.0) per 100,000 in 2050. Under the most pessimistic scenario, we estimated there would be 1,168,000 (95%CI: 983,000–1,324,000) additional tuberculosis cases and 95,000 (95%CI: 83,000–106,000) additional tuberculosis deaths in high-income countries over 2025-2050, with incidence of 11.5 (95%CI: 9.5–14.0) per 100,000 in 2050. The United States, United Kingdom, Germany, France, and Italy were projected to be the most affected high-income countries.

**Interpretation:** For high-income countries, the future risks of tuberculosis incidence and mortality could vary by as much as 5-times depending on the success or failure of tuberculosis control in high-burden settings, fundamentally shaping the strategies required to prevent, detect and treat tuberculosis in these settings.

## Introduction

With the waning of the COVID-19 pandemic, tuberculosis (TB) is again the leading global cause of infectious disease deaths—as it has been for most of the past century—and in 2024 caused 1.2 million deaths worldwide. A key feature of TB natural history is the delay between initial infection with *Mycobacterium tuberculosis* (*Mtb*, the causative agent of TB), and the development of symptomatic TB disease. With rapid disease progression this delay could be as short as 6 months, while for some individuals TB will be diagnosed decades after initial infection.^1^ This delay sets TB apart from other respiratory infectious diseases, and is particularly consequential for high-income countries that have achieved ongoing declines in *Mtb* transmission rates. For these countries, the majority of newly diagnosed TB cases may be due to infections acquired many years previously.^2,3^

International migration also plays a major role in shaping TB trends, as many migrant groups face elevated risks of TB.^4^ In 2024 there were an estimated 304 million people living in a country other than their birth, representing 3.7% of the global population, a substantial increase from an estimated 173 million in 2000.^5^ In coming decades the total number of migrants is forecast to continue increasing.^6^ International migration tends to move along known “corridors” shaped by economic, geographic, demographic, political, and cultural factors, typically from lower-income to higher-income countries, such as from Central America through Mexico to the United States, from Syria to Turkey, and from India to the United Arab Emirates. Leading destinations include Europe (94 million, 31% of 2024 total), North America (61 million, 20% of total), and the region of Northern Africa and Western Asia (54 million, 18% of total).^5^

A substantial proportion of new TB cases in high-income countries arise among individuals who have migrated from another country. This proportion varies widely across countries, representing over three-quarters of all TB cases in Australia, Germany, the Netherlands, Switzerland, the United Kingdom, and the United States, to less than one-quarter in Japan, Poland, Romania, South Korea, and Uruguay.^7^ However, the fraction of TB cases among foreign-born individuals is rising across almost all high-income countries. Future projections of TB incidence in the United States suggest a continuation of this trend,^8^ and similar trends may be expected for demographically similar high-income countries. Due to low transmission rates in high-income countries, most TB cases among foreign-born residents will be due to reactivation of *Mtb* infection acquired prior to migration.^9,10^ These dynamics have shaped strategies to address TB in high-income countries, with a major focus on interventions designed to identify and offer preventive treatment for *Mtb* infection among new migrants and foreign-born residents.^11^

The risks of TB faced by foreign-born individuals will be determined by their cumulative TB exposure prior to migration, as well as by preventive interventions received in their new country. Prior mathematical modeling of TB in the United States has illustrated how changes in TB exposure among future migrants can affect population-level TB risks, and the potential benefits within the United States of combined interventions that support TB control in high-burden countries while simultaneously strengthening TB prevention domestically.^12,13^ This suggests that high-income countries may benefit substantially from improved TB detection, treatment, and prevention in high-burden countries.

The future course of TB in low and middle-income countries is highly uncertain. The World Health Organization has established an ambitious plan to substantially reduce global TB incidence and mortality by 2035,^14^ supported by major political declarations at the UN General Assembly and by highly affected countries.^15,16^ Optimism for this plan has been bolstered by new developments in TB detection and treatment,^17–19^ and the possibility that new TB vaccines may soon be available.^20,21^ However, many countries with a high burden of TB rely heavily on donor assistance to provide TB services.^22^ With the elimination of USAID in early 2025, and reduced funding commitments by key international donors, it is possible that programs designed to combat TB in high-burden countries will shrink in coming years. Analyses that have forecast the potential implications of these changes have found that achieving global TB elimination targets could save millions of lives in highly affected countries.^23^ In contrast, the TB services disruptions caused by reduced health aid would cause major increases in TB incidence and mortality, setting back TB elimination efforts by several decades.^24,25^

Given these dynamics, the future course of TB epidemics in high-burden settings could have major implications for high-income countries, affecting TB risks within these countries, and the interventions and resources that will be required to address them. In this study we investigated how the future risks of tuberculosis in high-income countries will change depending on the success or failure of efforts to combat tuberculosis in high-burden settings. We did so by defining scenarios representing different levels of optimism regarding tuberculosis control in high-burden settings, and using calibrated mathematical models to predict how these scenarios would change *Mtb* infection carriage among future migrants, and thereby change tuberculosis incidence and deaths in high-income countries. The analysis considered the majority of countries involved in migration-linked TB—either as origin countries or destination countries—and projected outcomes until 2050.

## Methods

### Countries included in the analysis

#### Origin countries

These were selected to include countries with a high current burden of TB. To do so, we selected all countries listed by the WHO as having a high burden of TB, TB/HIV, and/or multidrug-resistant TB, comprising a total of 49 countries (**Table S1**).^26^ Countries were included in these lists if they were among the top 20 countries by estimated absolute number of incident cases or among the top 10 countries by incidence rate in 2020 for TB, TB among people with HIV, or multidrug-resistant TB, respectively. The majority of selected countries were low or middle-income countries according to World Bank country classifications (all except Russia, designated high-income in 2024), and represented 90% of global TB incidence in 2024.^27^

#### Destination countries

These were selected to include 60 World Bank-designated high-income countries and autonomous territories (**Table S2**).^28^ We excluded Russia due to its recent transition to high-income country status, and its inclusion in the list of origin countries with high TB burden. We also excluded 25 low-population countries. The final set of countries represented 96% of all foreign-born TB cases reported by high-income countries for 2024.^7^

### Scenarios

We created several scenarios for the future trajectory of TB epidemiology in countries with a high burden of TB:

#### Scenario 1, Continuation of pre-2025 trends (base-case)

This scenario assumed that pre-2025 trends in TB epidemiology would continue in each high-burden country, based on the annual percentage change in WHO-estimated TB incidence rates averaged over the past 10 years.^27^

#### Scenario 2, USAID and Global Fund support ended in 2025

This scenario assumed that two major sources of international support to TB programs—U.S. bilateral aid provided through USAID, and funding from the multilateral Global Fund to Fight AIDS, Tuberculosis, and Malaria (Global Fund)—would be removed from each high-burden country from 2025 onwards. To operationalize this scenario we extracted WHO data on TB program expenditures for each origin country in the most recent available year, stratified by funding source (domestic, US bilateral, Global Fund, other international).^22^ We calculated the share of program funding provided by either USAID or Global Fund, and assumed that removal of these funding sources would reduce overall program funding proportionally. We used an approach developed in an earlier analysis to represent the effect that funding cuts would have on TB services, operationalized as reductions in treatment access (proportion of individuals developing tuberculosis disease that are diagnosed and treated). We assumed that funding reductions would produce a less-than-proportional reduction in treatment access, due to reallocation of resources to highest priority interventions, improvements in technical efficiency, and other efforts to counteract the disruption caused by funding cuts.^25^ We operationalized this as an adjustment ratio of 0.50, such that a 50% funding reduction would reduce treatment access by 25%. These assumptions were applied separately for each country as modifications to the base-case scenario, from 2025 onwards. This represented the most pessimistic scenario analyzed.

#### Scenario 3, USAID support ended in 2025

This scenario assumed that US bilateral aid provided through USAID would be cut from 2025 onwards (following the shuttering of USAID in early 2025), but that all other sources of TB program funding would be unaffected. We used the same approach as the “*USAID and Global Fund support ended*” scenario to operationalize these changes, applied as modifications to the base-case scenario for each country from 2025 onwards.

#### Scenario 4, Cautious optimism

This scenario assumed a 3-percentage point acceleration in the rate of decline in TB incidence in each high-burden country from 2025 onwards, achieved through expanded treatment access for TB disease. This acceleration was based on the observed variation in TB incidence trends within the set of modelled countries over the past 10 years,^27^ representing the difference between the median (50^th^ percentile) and the upper quartile (75^th^ percentile) of the annual percentage decline in TB incidence. This scenario was designed to represent improvements in TB control within the range of recent programmatic experience.

#### Scenario 5, Achievement of End TB Strategy targets

This scenario assumed a rapid acceleration in TB care and prevention to achieve the epidemiological targets described in the WHO’s End TB Strategy,^14^ including an 80% reduction in TB incidence rates and 95% reduction in TB mortality from 2015 levels by 2030. Under plans developed to meet End TB Strategy targets, accelerated TB control is to be achieved through large-scale expansion of existing interventions and deployment of new technologies.^29^ We operationalized these changes through expanded access to TB disease treatment, scale-up of preventive treatment for individuals with *Mtb* infection, and the introduction of a new TB vaccine in 2030. This represented the most optimistic scenario analyzed.

### Recent trends in destination countries

We extracted WHO data on the number of foreign-born and total individuals diagnosed with TB for each destination country. We calculated the number of TB diagnoses for native-born individuals by subtracting foreign-born diagnoses from total diagnoses.^7^ We analyzed these data to report recent trends in foreign-born and native-born TB cases within the 57 destination countries, individually and overall.

### Modeling of future trends

We projected future TB incidence and mortality trends in destination countries under each scenario, using a modelling approach. **Figure S1** provides a schematic of data sources, analyses, and outcomes.

#### Simulation of TB epidemiology in high TB-burden countries

We adopted an existing transmission dynamic model developed in prior studies,^12,25^ and modified it to represent the five scenarios addressed in this analysis. For each of the 59 origin countries the model was parameterized with demographic estimates reported by the UN Population Division,^30^ and calibrated to match WHO TB incidence estimates for 2000-2023 as well as published estimates of *Mtb* infection prevalence.^27,31^ In the model, the prevalence and recency of *Mtb* infection in a given age group and year reflects infectious exposure over the individual’s lifetime, based on the time-series of transmission rates within the country. Results from the fitted models were used to parameterize *Mtb* infection carriage among migrants leaving each origin country, as described below.

#### International migration flows from origin to destination countries

Estimated migration volumes were drawn from published probabilistic forecasts.^6^ These forecasts report estimates of international migration between each origin-destination country pair in 5-year periods, covering 2020 to 2045. We used linear interpolation to transform these five-year forecasts into single-year values. The ages of individuals in migrating cohorts were based on the age distribution of recently-arrived foreign-born individuals included in American Community Survey (ACS) microdata for 2017-2021 (**Figure S2**).^32^

#### Projection of TB incidence and mortality in destination countries

We projected total population TB cases and deaths in each destination country for 2025-2050. *Mtb* infection prevalence in migrating cohorts was based on the outcomes of the origin country models, stratified by year, age group, and scenario. As migrants may have higher or lower rates of *Mtb* infection compared to their origin country population, we calibrated *Mtb* infection prevalence in migrating cohorts to reproduce reported numbers of total foreign-born TB notifications in each destination country for 2015-2023,^7^ adjusted for country-specific rates of under-detection estimated by WHO.^27^ Future TB cases and deaths for each migrating cohort were simulated using a Markov model that tracked TB natural history, incidence, and mortality based on *Mtb* infection rates at the point of migration and general population background mortality rates for each destination country.^30^ Natural history assumptions followed approaches used in past analyses,^33^ and matched the model structures used in origin country models. Case fatality rates for individuals developing TB disease were calculated from Global Burden of Disease Study estimates, stratified by destination country and age group.^34^ We did not account for ongoing post-migration TB exposure for migrants, based on low transmission rates in destination countries compared to origin countries.

To understand how each scenario would affect overall population-level TB incidence we forecast trends in TB case rates among native-born populations, and added these to simulated foreign-born TB cases to calculate total cases. To do so, we fit log-linear time trends to the series of native-born TB cases for each destination country.^7^ Future time trends were estimated using a negative binomial regression model and log link function, with country random effects for intercept and time trend. This approach assumed no difference in the number of native-born TB cases between scenarios, consistent with published molecular epidemiological studies showing minimal *Mtb* transmission between foreign-born and native-born populations.^35^

### Statistical analysis

We used second-order Monte Carlo simulation to propagate uncertainty in analytic inputs and calculate measures of precision around study outcomes.^36^ Probability distributions were defined to represent uncertainty in major analytic inputs, including TB natural history parameters, migration volumes, migrant age distribution, and native-born TB time trends. These probability distributions were created as closed-form functions parameterized to reproduce the mean and reported uncertainty intervals of existing estimates (**Table S3**). For future time trends in TB among native-born populations, we simulated incidence trajectories using the point estimates and variance-covariance matrix of the fitted regression models. To allow for uncertainty in age at migration we reweighted the existing age distribution to allow for a +/- 5-year variation in mean age for migrating cohorts. We randomly sampled 1000 sets of values for each input using a Latin Hypercube sampling approach and re-ran the analysis using each of these parameter sets, producing 1000 estimates for each study outcome. Finally, we calculated point estimates and 95% credible intervals as the mean and 2.5^th^ – 97.5^th^ percentiles of the distribution of results for each outcome of interest. All analyses were conducted in R version 4.2.2. Origin country models were programmed in C++ via the Rcpp package (v1.0.9).^37,38^

### Validation

We validated study estimates of foreign-born and native-born TB incidence in each destination country against empirical data reported for 2024,^7^ which were not used for model fitting.

### Human subject protections

This study does not constitute human subjects research. All analyses were conducted with aggregated publicly-available data, including TB notification data reported to WHO by country governments,^7^ and ACS public-use microdata.^32^

## Results

### Recent trends in destination countries

**Figure 1** shows trends in TB diagnoses in 60 high-income destination countries from 2009 to 2024. Over this period 26% (558,713 of 2,112,025) of all TB diagnoses in these countries were among foreign-born individuals. The fraction of TB cases among foreign-born individuals increased in 13 out of 15 years, from 20% in 2009 to 39% in 2024. The overall fraction of foreign-born TB cases varied widely across countries (**Figure S3**), with an interquartile range (IQR) of 13% to 73%. However, time trends within each country were notably consistent, with the number of native-born TB cases declining more rapidly than foreign-born TB cases (**Figure S4**). Across the 60 destination countries the median annual percent change in foreign-born TB cases was 0.2%, (IQR: -3.3% to 4.2%), while for native-born populations the median annual percent change was -5.2% (IQR: -6.9% to -1.4%).

**Figure 1:**
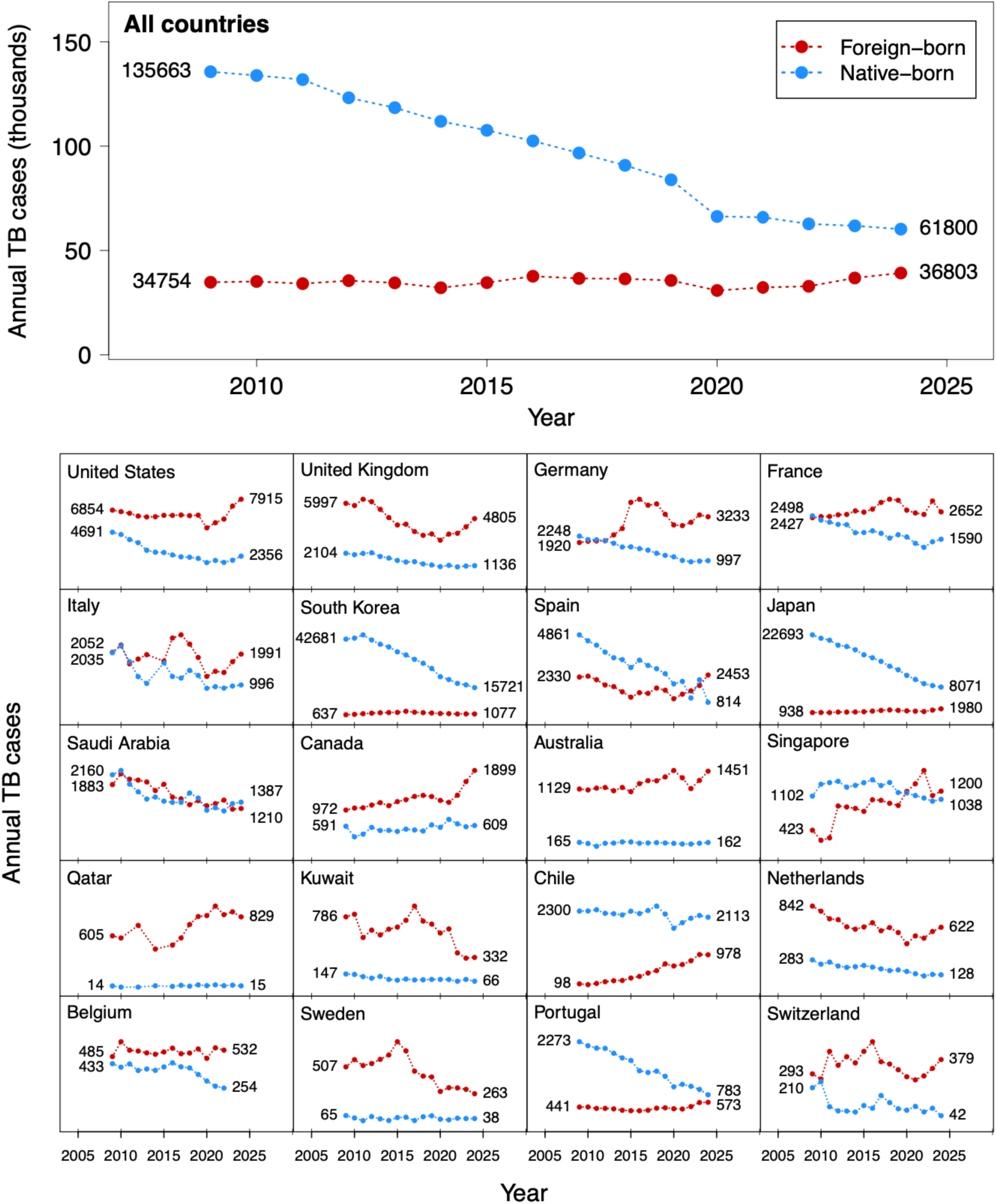
Time trends in TB among foreign-born and native-born individuals over 2009-2024, overall and for selected countries. Countries shown include top 20 countries in terms of TB cases among foreign-born individuals over 2014-2024. Data for all destinations countries shown in **Figure S4**.

### Validation against 2024 data

We validated short-term model predictions by comparing study estimates of TB incidence in 2024 against data reported to WHO for the same year, which were not used for model fitting. These results are shown in **Figure S5**, demonstrating close agreement of study estimates and empirical data, for both foreign-born and native born populations.

### Base-case projections of future migrant TB cases

We forecast future trends in TB among migrant populations, assuming continuation of pre-2025 TB trends in migrant origin countries. Under this scenario, we predicted a total 1.29 million (95% credible interval (CI): 1.01–1.75) foreign-born individuals would develop TB between 2025 and 2050 in the 60 destination countries included in the analysis, with 483,000 (95%CI: 370,000–666,000) of these during the 2025-2035 period. Most of these foreign-born TB cases were predicted for the European Region, with 689,000 (95%CI: 560,000–896,000) cases predicted for 2025-2050, 53% (95%CI: 50–57) of the total. A total of 299,000 (95%CI: 212,000–436,000) foreign-born TB cases were predicted for the Americas Region (23% (95%CI: 21–25) of total), 218,000 (95%CI: 163,000–301,000) for the Western Pacific Region (17% (95%CI: 16–18) of total), and 87,000 (95%CI: 58,000–136,000) for the Eastern Mediterranean Region (7% (95%CI: 5–8) of total). **Figure 2** shows predicted TB cases for the top 20 destination countries, with the greatest number of cases predicted for the United States (232,000 cases, 95%CI: 163,000–343,000), followed by the United Kingdom (158,000 cases, 95%CI: 124,000–214,000), France (153,000 cases, 95%CI: 120,000–190,000), and Germany (103,000 cases, 95%CI: 81,000–136,000). **Figure S6** reports results for all 60 destination countries. The annual number of foreign-born TB cases was predicted to increase progressively over the projection period, from 41,000 (95%CI: 31,000–58,000) in 2025 to 59,000 (95%CI: 45,000–80,000) in 2050, representing an average annual increase of 1.47% (95%CI: 0.99–2.05). The number of TB deaths for migrant populations followed similar trends, with 112,000 (95%CI: 82,000–161,000) total migrant TB deaths predicted for the 2025-2050 period (average case fatality 8.6%, 95%CI: 7.5–9.8).

**Figure 2:**
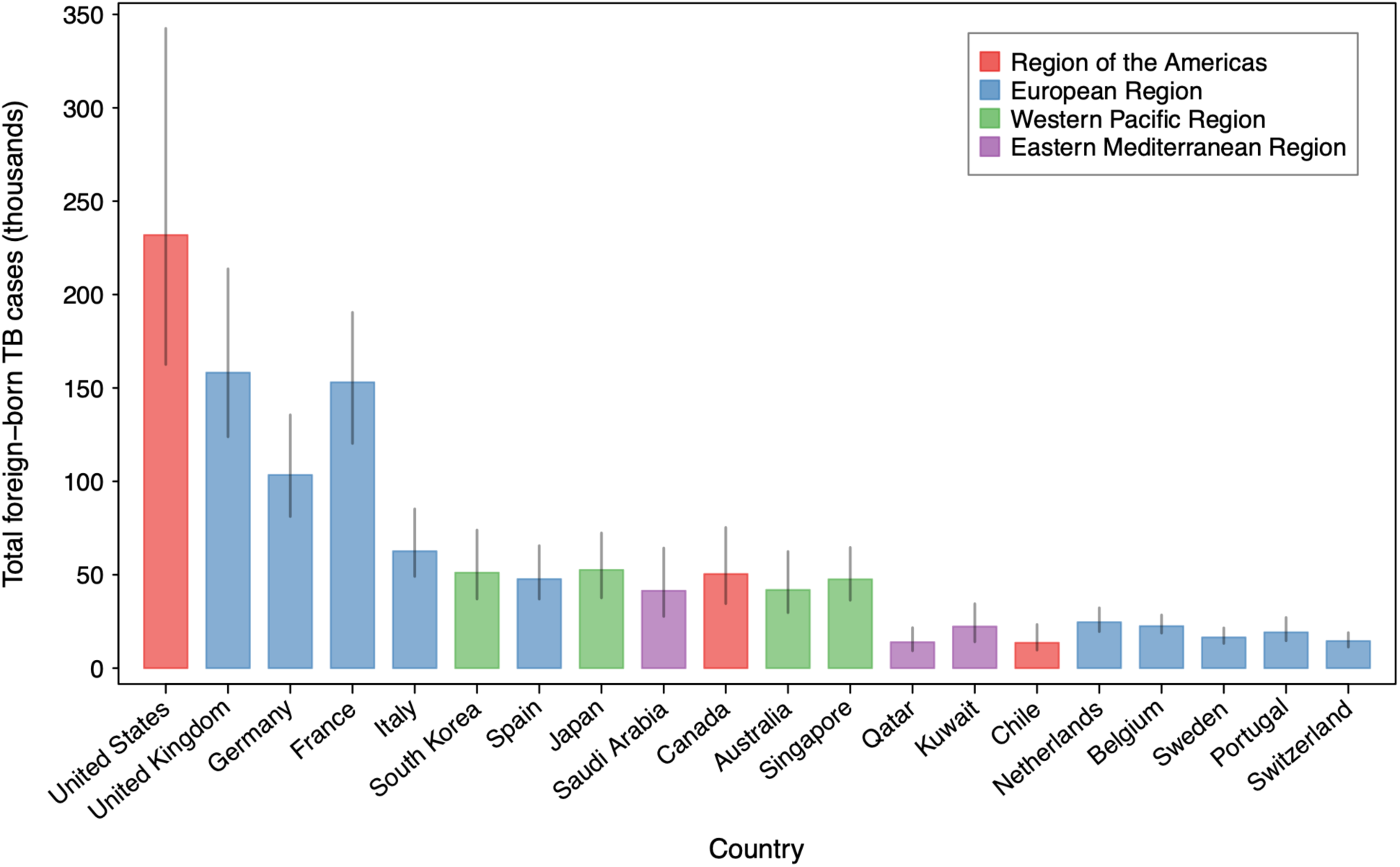
Projected numbers of foreign-born individuals developing TB over 2025-2050 for the top 20 destination countries, under the base-case scenario. Vertical lines represent 95% credible intervals. Countries ordered by total foreign-born TB cases reported for 2014-2024.

### Migrant TB cases under alternative scenarios

**Table 1** reports predicted future numbers of TB cases and deaths among foreign-born individuals in the 60 high-income destination countries for the five scenarios representing different TB trends in migrants’ origin countries. Under the most pessimistic scenario, representing the permanent termination of health aid provided by USAID and the Global Fund, foreign-born TB cases were predicted to total 2,462,000 (95%CI: 2,058,000–3,040,000) over 2025-2050, a 92% (95%CI: 69–112) increase compared to the base-case. Under the most optimistic scenario, which envisaged rapid reductions in TB incidence and transmission for each origin country consistent with the WHO’s End TB Strategy, foreign-born TB cases in destination countries were predicted to total 509,000 (95%CI: 315,000–820,000), a 61% (95%CI: 52–69) reduction compared to the base-case. Even more modest improvements in TB control in origin countries were projected to produce meaningful changes in destination countries, with a 3% acceleration in the rate of incidence reductions in origin countries (‘Cautious optimism’ scenario) projected to reduce total foreign-born TB cases over 2025-2050 by 373,000 (313,000–427,000) compared to the base-case, a 29% (95%CI: 23–35) reduction.

**Table 1:** Projected numbers of TB cases and deaths among foreign-born individuals in modelled destination countries between 2025 and 2050 under different scenarios. Values in parentheses represent 95% uncertainty intervals

| Scenario | TB cases |  | TB deaths |  |
| --- | --- | --- | --- | --- |
|  | 2025-2035 | 2025-2050 | 2025-2035 | 2025-2050 |
| **Total TB cases and deaths among foreign-born individuals (thousands)** |  |  |  |  |
| Continuation of pre-2025 trends (base-case) | 483<br>(370, 666) | 1,294<br>(1,008, 1,753) | 42<br>(30, 62) | 112<br>(82, 161) |
| USAID and Global Fund support ended in 2025 | 716<br>(589, 914) | 2,462<br>(2,058, 3,040) | 61<br>(47, 82) | 207<br>(170, 263) |
| USAID support ended in 2025 | 504<br>(392, 686) | 1,394<br>(1,095, 1,864) | 44<br>(32, 64) | 120<br>(90, 170) |
| Cautious optimism | 402<br>(292, 578) | 921<br>(663, 1,352) | 36<br>(24, 56) | 83<br>(55, 131) |
| Achieve End TB Strategy targets | 340<br>(232, 511) | 509<br>(315, 820) | 31<br>(19, 51) | 49<br>(27, 86) |
| **Absolute difference compared to base-case scenario (thousands)** |  |  |  |  |
| USAID and Global Fund support ended in 2025 | 233<br>(180, 276) | 1,168<br>(983, 1,324) | 19<br>(15, 23) | 95<br>(83, 106) |
| USAID support ended in 2025 | 21<br>(16, 25) | 100<br>(82, 116) | 2<br>(1, 2) | 8<br>(7, 10) |
| Cautious optimism | -80<br>(-95, -63) | -373<br>(-427, -313) | -6<br>(-7, -5) | -29<br>(-33, -25) |
| Achieve End TB Strategy targets | -143<br>(-170, -110) | -785<br>(-950, -647) | -11<br>(-13, -9) | -63<br>(-78, -52) |
| **Percentage change compared to base-case scenario (%)** |  |  |  |  |
| USAID and Global Fund support ended in 2025 | 49<br>(33, 65) | 92<br>(69, 112) | 47<br>(29, 65) | 88<br>(61, 114) |
| USAID support ended in 2025 | 4<br>(3, 6) | 8<br>(6, 10) | 4<br>(3, 6) | 8<br>(5, 10) |
| Cautious optimism | -17<br>(-22, -12) | -29<br>(-35, -23) | -15<br>(-21, -10) | -27<br>(-34, -19) |
| Achieve End TB Strategy targets | -30<br>(-38, -21) | -61<br>(-69, -52) | -27<br>(-37, -18) | -57<br>(-67, -47) |

**Table 2** reports incremental numbers of TB cases under each scenario as compared to the base-case, for each of the top 20 destination countries. For the United States, the termination of USAID and Global Fund support in 2025 was projected to produce 228,000 (95%CI: 180,000–288,000) additional TB cases over 2025-2050, while achievement of the End TB Strategy targets in origin countries would reduce TB cases over the same period by 139,000 (95%CI: 106,000–183,000). Using the difference in projected TB cases between most optimistic and pessimistic scenarios as an indicator of a country’s exposure to future changes in TB trends in origin countries, the top 10 countries (in order: United States, France, United Kingdom, Germany, Italy, Canada, Belgium, Spain, South Korea, and Australia) accounted for 76% (95%CI: 75–78) of the total for all destination countries, and the top 4 alone accounted for 56% (95%CI: 54–58). **Table S4** and **Table S5** report country-specific estimates of incremental TB cases and deaths, respectively, for each scenario compared to the base-case. **Table S6** reports percentage changes in TB cases compared to the base-case.

**Table 2:** Incremental numbers of TB cases among foreign-born individuals projected for 2025-2050 in top twenty destination countries under different scenarios, as compared to base-case. Table shows results for top 20 countries by total foreign-born TB notifications over 2014-2024. Countries ordered by total foreign-born TB notifications over 2014-2024. Positive values indicate additional TB cases as compared to the base-case. Negative values indicate fewer TB cases as compared to the base-case. Values in parentheses represent 95% uncertainty intervals.

| Country | Incremental TB cases (thousands), by scenario |  |  |  |
| --- | --- | --- | --- | --- |
|  | USAID and Global Fund support ended in 2025 | USAID support ended in 2025 | Cautious optimism | Achieve End TB Strategy targets |
| United States | 228 (180, 288) | 11 (8, 13) | -67 (-86, -53) | -139 (-183, -106) |
| United Kingdom | 154 (119, 184) | 7 (6, 10) | -46 (-53, -39) | -96 (-118, -80) |
| Germany | 151 (121, 189) | 16 (13, 18) | -30 (-35, -24) | -62 (-75, -50) |
| France | 168 (119, 218) | 20 (14, 27) | -44 (-52, -35) | -100 (-121, -77) |
| Italy | 51 (39, 62) | 3 (3, 4) | -18 (-20, -15) | -36 (-43, -30) |
| South Korea | 23 (16, 29) | 4 (3, 5) | -15 (-22, -11) | -32 (-50, -23) |
| Spain | 33 (27, 39) | 4 (3, 6) | -14 (-18, -11) | -27 (-35, -21) |
| Japan | 21 (16, 26) | 3 (2, 4) | -16 (-20, -11) | -31 (-40, -22) |
| Saudi Arabia | 22 (18, 27) | 4 (3, 5) | -11 (-13, -9) | -24 (-32, -18) |
| Canada | 41 (33, 48) | 2 (2, 3) | -14 (-18, -11) | -31 (-42, -23) |
| Australia | 27 (22, 34) | 2 (2, 3) | -12 (-15, -10) | -25 (-35, -19) |
| Singapore | 18 (13, 23) | 4 (3, 5) | -14 (-18, -12) | -32 (-41, -25) |
| Qatar | 4 (3, 6) | 1 (1, 1) | -3 (-4, -3) | -7 (-10, -5) |
| Kuwait | 10 (7, 14) | 1 (1, 1) | -6 (-11, -4) | -13 (-23, -8) |
| Chile | 10 (5, 19) | 3 (1, 5) | -4 (-7, -3) | -8 (-14, -5) |
| Netherlands | 23 (18, 29) | 3 (2, 4) | -7 (-9, -6) | -16 (-20, -12) |
| Belgium | 49 (34, 74) | 4 (3, 6) | -7 (-8, -6) | -14 (-17, -12) |
| Sweden | 18 (15, 21) | 1 (1, 1) | -5 (-6, -4) | -10 (-13, -8) |
| Portugal | 7 (6, 9) | 0 (0, 0) | -6 (-7, -5) | -10 (-14, -8) |
| Switzerland | 14 (11, 23) | 1 (1, 1) | -4 (-5, -4) | -9 (-11, -7) |

### Trends in general population incidence rates

We combined projections of TB among foreign-born populations with statistical projections of TB among native-born populations to understand possible trends in general population TB rates. **Figure 3** shows projections of future general population TB incidence for the 60 destination countries under each scenario. Under the base-case scenario, total TB incidence is projected to be 2,266,000 (95%CI: 1,938,000–2,744,000) over 2025-2050, with 57% (95%CI: 50–65) of total cases among foreign-born individuals. TB incidence is projected to decline slowly from 104,000 (95%CI: 93,000–121,000) cases per year in 2025 and then plateau after 2035, reaching 82,000 (95%CI: 67,000–104,000) cases per year in 2050. In contrast, ongoing incidence reductions are projected for scenarios that assumed accelerated TB control in origin countries, with 2050 incidence estimated as 58,000 (95%CI: 46,000–76,000) TB cases for the Cautious Optimism scenario, and 29,000 (95%CI: 23,000–37,000) for the scenario assuming achievement of the End TB Strategy targets. For the most pessimistic scenario, assuming termination of USAID and Global Fund health aid, TB incidence increases through most of the projection period, to 144,000 cases (95%CI: 119,000–175,000) in 2050. **Table S7** reports the percentage of TB cases among foreign-born individuals in 2050 for each scenario and destination country. Summing across all 60 destination countries, this proportion ranged from 21% (95%CI: 11, 35) under the most optimistic scenario to 84% (905%CI: 79, 88) under the most pessimistic.

**Figure 3:**
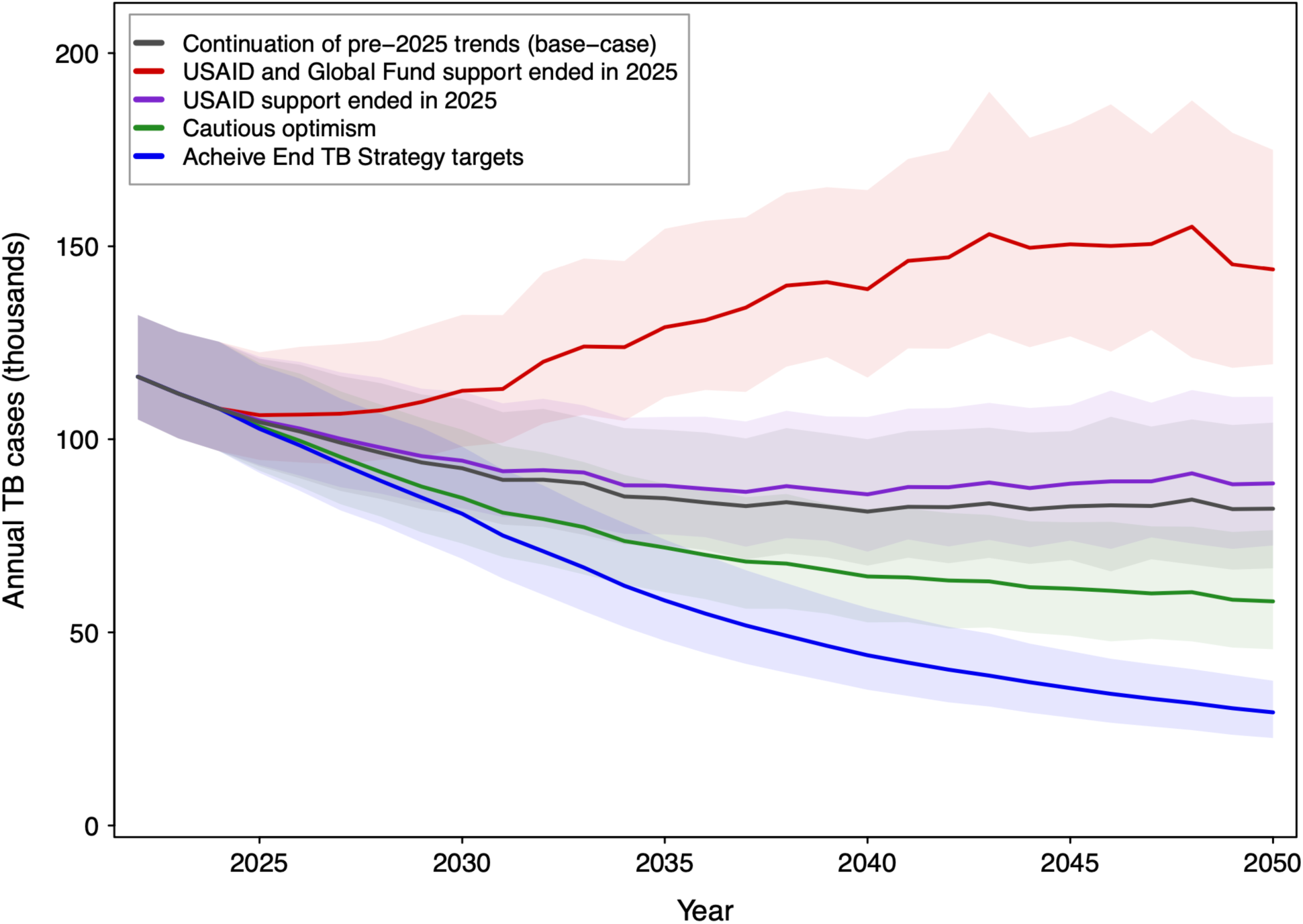
Projected annual numbers of TB cases in destination countries between 2025 and 2050 under different scenarios. Lines represent point estimates. Shaded regions represent 95% uncertainty intervals. TB cases include both native- and foreign-born individuals.

**Table 3** reports general population TB incidence rates predicted for 2050 in the top twenty destination countries under each scenario, compared to different TB control targets. Compared to the WHO’s pre-elimination target of <1 annual case per 100,000, four of these countries are predicted to achieve this threshold by 2050 under the most optimistic scenario, and none under the other scenarios. For a more permissive threshold of <5 cases per 100,000, 17 out of 20 countries would achieve this by 2050 under the most optimistic scenario, compared to 2 of 20 in the base-case, and 1 of 20 in the most pessimistic scenario. **Table S8** reports results for all destination countries.

**Table 3:** Projected TB incidence rate in general population for 2050 in top twenty destination countries under different scenarios. Countries ordered by total foreign-born TB notifications over 2014-2024. Values in parentheses represent 95% uncertainty intervals. Cells coded by attainment of TB incidence thresholds by 2050: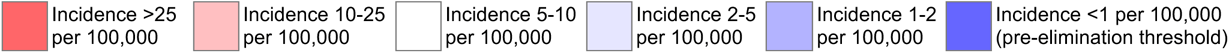

| Country | TB incidence rate in 2050 (cases per 100,000 person-years) |  |  |  |  |
| --- | --- | --- | --- | --- | --- |
|  | USAID and Global Fund support ended in 2025 | USAID support ended in 2025 | Continuation of pre-2025 trends (base-case) | Cautious optimism | Achieve End TB Strategy targets |
| All countries | 11.5 (9.5, 14.0) | 7.1 (5.8, 8.9) | 6.5 (5.3, 8.3) | 4.6 (3.6, 6.1) | 2.3 (1.8, 3.0) |
| United States | 6.4 (4.2, 9.1) | 3.2 (2.1, 4.9) | 3.0 (1.9, 4.7) | 1.8 (1.1, 3.0) | 0.5 (0.2, 0.8) |
| United Kingdom | 20.3 (15.3, 25.5) | 10.1 (7.5, 14.0) | 9.5 (6.8, 13.3) | 5.7 (3.8, 8.8) | 1.4 (0.7, 2.5) |
| Germany | 14.3 (11.0, 18.9) | 6.9 (5.4, 8.9) | 5.7 (4.4, 7.5) | 3.5 (2.5, 4.9) | 0.9 (0.5, 1.6) |
| France | 25.1 (15.2, 37.2) | 14.7 (9.5, 23.4) | 12.6 (8.4, 22.0) | 8.5 (5.7, 14.8) | 2.1 (1.2, 3.8) |
| Italy | 11.5 (6.4, 22.1) | 6.9 (4.0, 16.5) | 6.5 (3.7, 15.0) | 4.1 (2.3, 8.1) | 1.4 (0.7, 2.6) |
| South Korea | 12.4 (7.2, 20.1) | 10.5 (5.9, 18.0) | 10.0 (5.6, 17.4) | 8.1 (4.2, 15.1) | 5.8 (2.3, 12.3) |
| Spain | 10.1 (6.5, 14.4) | 6.3 (4.3, 8.5) | 5.5 (3.9, 7.5) | 3.6 (2.5, 5.2) | 1.5 (0.8, 2.5) |
| Japan | 4.6 (2.8, 6.9) | 3.8 (2.3, 5.9) | 3.6 (2.2, 5.7) | 2.7 (1.5, 4.6) | 1.7 (0.8, 3.5) |
| Saudi Arabia | 8.0 (5.1, 12.6) | 5.7 (3.4, 9.2) | 5.3 (3.1, 8.1) | 4.0 (2.1, 6.5) | 2.3 (1.0, 4.5) |
| Canada | 14.4 (9.3, 20.4) | 9.4 (5.3, 14.8) | 9.1 (5.1, 14.3) | 6.7 (3.6, 11.7) | 4.0 (1.7, 8.3) |
| Australia | 11.0 (7.9, 15.2) | 6.7 (4.4, 9.7) | 6.3 (4.1, 9.2) | 3.8 (2.3, 6.0) | 0.9 (0.4, 1.6) |
| Singapore | 69.3 (42.0, 114.1) | 55.7 (31.0, 96.6) | 51.2 (28.6, 87.6) | 33.9 (18.8, 55.4) | 13.4 (5.6, 28.5) |
| Qatar | 22.0 (13.8, 39.1) | 16.5 (9.8, 29.9) | 15.3 (8.9, 27.9) | 9.9 (5.3, 17.6) | 3.5 (1.1, 9.6) |
| Kuwait | 27.2 (13.2, 54.8) | 17.9 (8.2, 39.6) | 17.1 (7.8, 39.1) | 10.3 (4.4, 22.4) | 2.2 (0.9, 4.3) |
| Chile | 16.2 (7.2, 30.1) | 13.6 (6.3, 26.2) | 12.6 (5.8, 25.4) | 11.4 (5.0, 24.5) | 10.2 (3.8, 23.4) |
| Netherlands | 12.0 (9.0, 18.4) | 6.7 (4.6, 10.6) | 5.8 (3.9, 9.1) | 3.6 (2.3, 5.7) | 0.7 (0.4, 1.3) |
| Belgium | 30.0 (19.0, 46.5) | 12.4 (8.5, 17.6) | 10.0 (7.0, 13.7) | 6.3 (4.4, 9.2) | 1.7 (0.8, 3.2) |
| Sweden | 15.1 (10.4, 25.8) | 7.8 (5.1, 13.4) | 7.2 (4.7, 12.8) | 4.6 (2.9, 7.8) | 1.4 (0.7, 2.5) |
| Portugal | 15.6 (10.1, 27.0) | 11.8 (7.2, 21.0) | 11.6 (7.0, 20.9) | 7.7 (4.4, 13.2) | 3.9 (2.0, 7.0) |
| Switzerland | 15.5 (11.6, 20.2) | 8.6 (6.2, 11.8) | 8.0 (5.7, 11.0) | 5.3 (3.6, 7.9) | 2.1 (1.1, 4.0) |

## Discussion

In this study we examined how the future risks of tuberculosis in high-income countries could change depending on the success or failure of efforts to combat tuberculosis in high-burden settings. To do so, we modeled patterns of migration-linked TB between 49 high-burden origin countries and 60 high-income destination countries, using a system of mathematical models fit to available epidemiological and demographic data. Assuming that pre-2025 trends in origin countries continue, we projected approximately 1.3 million foreign-born individuals in high-income destination countries will develop TB over 2025-2050, with annual case totals increasing slowly from current levels. In contrast, alternative scenarios that assumed rapid acceleration or deterioration of TB control in origin countries produced major consequences for TB in high income countries, with total 2025-2050 migrant TB cases under these alternative scenarios varying from 39% to 192% of the base-case projection. General population incidence rates in high-income countries also varied widely under these different scenarios, with an average 2050 incidence rate of 2 per 100,000 under the most optimistic scenario, and 11 per 100,000 under the most pessimistic scenario. These results highlight the central role of migration-linked TB in shaping TB epidemiology in high-income countries. The United States, United Kingdom, Germany, France, and Italy were the countries projected to experience the greatest absolute impact, but some smaller countries had similar relative effects. Projected changes in general population incidence rates were smaller for countries with larger numbers of TB cases among native-born populations, and lower migration rates.

The base-case projections of this analysis are consistent with prior studies that have highlighted the elevated TB risks faced by individuals who have migrated from high- to low-burden settings, and the major role that migration-linked TB plays in TB epidemiology in many high-income countries.^4,10,39,40^ These have motivated TB prevention policies in high-income countries that address the elevated risk faced by foreign-born individuals, including screening for active TB disease at the time of entry and provision of TB preventive treatment (TPT) to new or established migrants.^9,41,42^ Our base-case results suggest that these interventions will be increasingly important, given the growing share of TB projected for migrant populations.

Across the scenarios examined in this analysis, future TB incidence rates for most high-income countries were sensitive to TB trends in high-burden counties. The magnitude of these differences can be benchmarked against the potential impact of currently available interventions. Studies conducted in the United Kingdom have demonstrated reduced TB incidence for interventions that screen migrants for TB disease (either before entry or after arrival) and/or offer TPT, reporting 20-35% reductions in subsequent TB incidence rates for individuals receiving one of these interventions compared to those that did not.^43–45^ These health benefits are valuable, and TB screening and prevention for migrants has been found to be cost-effective in a number of studies.^46,47^ However, those who receive these interventions still experience elevated TB incidence compared to native-born populations.^48^ Moreover, these programs face difficulties in reaching high population coverage. For TPT, substantial losses have been documented at each step of the treatment cascade (testing acceptance, treatment initiation, treatment completion), such that in many studies only a small fraction of those eligible for this intervention completed it.^49–51^ Pre-entry TB disease screening can achieve much higher uptake rates when required for immigration, but will not apply to all migrants. For example, the United States requires TB screening for all individuals applying to become legal permanent residents,^52^ but this group represents only a small fraction of the total individuals entering the country each year, which also includes temporary residents, short-term visitors, and undocumented immigrants.^53^ Due to these factors, interventions provided during or after entry are an incomplete response to the TB risks faced by migrants.

In this context, efforts to strengthen TB services in high-burden countries could have substantial benefits for high-income countries. These benefits would be small compared to the great health benefits realized in high-burden countries,^54^ but would still greatly improve the prospects for TB elimination in low-burden settings, as well as delivering valuable health and economic benefits.^13^ Moreover, strengthening TB control efforts in high-burden settings has been shown to be complementary to strengthened domestic prevention services,^55^ with both included as priority action areas in TB policy frameworks developed for low-incidence settings.^56^ Changes in migration policy could also affect migration-linked TB by modifying short-term migration rates, yet we did not explore such policy scenarios in this analysis. While potential reductions in TB incidence in high-income might be seen as a justification for migration restrictions, the economic benefits of reduced TB incidence would be small compared to the foregone economic benefits of migration to destination countries, including increased net government revenues, faster economic growth, and greater productivity.^57,58^ Moreover, historical fluctuations in migration policy within high-income counties suggest that any new migration restrictions would likely have temporary consequences.^59^ The political environments that produce more restrictive migration policy may also harm healthcare access for migrants, potentially delaying treatment of TB disease and increasing mortality rates.

This study has several limitations. First, there is substantial uncertainty in many analytic inputs. In particular, future migration trends are difficult to predict, reflected in wide uncertainty intervals in the migration forecasts used for our analysis.^6^ Unanticipated geopolitical events or economic shifts could significantly alter migration trends, which in turn would affect patterns of migration-linked TB. Second, we did not explicitly model the TB prevention interventions currently provided in destination countries. As we calibrated the analysis to current TB reporting data for each destination country, the impact of these interventions will be implicitly factored into our estimates.^7^ However, future TB rates in destination countries would be lower if preventive services were to be expanded compared to current levels. Third, our analysis did not account for infection or reinfection of migrants after entry, an alternative explanation for elevated incidence rates many years after entry. While migrants likely face elevated infection risks compared to native-born populations (due to travel to origin countries, or contact with visitors from these countries), these risks would decline or increase proportional to the mechanisms considered in our study, such that their exclusion is unlikely to meaningfully affect results. Fourth, we did not consider transmission from foreign-born to native-born populations. Prior studies have found TB cases attributable to such transmission to be rare.^35^ If these cases were considered in the analysis, the incremental difference in TB cases between scenarios would be slightly greater. Fifth, we only considered a subset of countries in this analysis, either as destination or origin countries. Our selected origin countries included >90% of global TB incidence, and our destination countries >90% of foreign-born TB cases. However, estimated impacts would be larger if a more complete set of countries were included. Moreover, the approach of categorizing countries as origin and destination countries greatly simplifies the realities of global migration. Sixth, this analysis only considered a limited set of scenarios. Two of these scenarios—the most optimistic and pessimistic—represented extremes of what could happen in high-burden counties, and it is unlikely that actual outcomes will fall outside these bounds. Finally, this analysis only considered outcomes in high-income countries, yet the major impact of the scenarios assessed in this analysis will occur in low-income countries with a high burden of TB. These consequences have been quantified in other analyses,^24,25,54^ and in this study we focused on an outcome only previously estimated in single country case-studies.^13,55^

In summary, this study shows a close relationship between the relative success of tuberculosis control in high-burden settings and the future risks of tuberculosis incidence and mortality in high-income countries. Over coming decades, this relationship will fundamentally shape the strategies required to prevent, detect and treat tuberculosis in high-income countries.^60^

## Data Availability

All data produced in the present work are contained in the manuscript.

## SUPPLEMENT

**Table S1:**
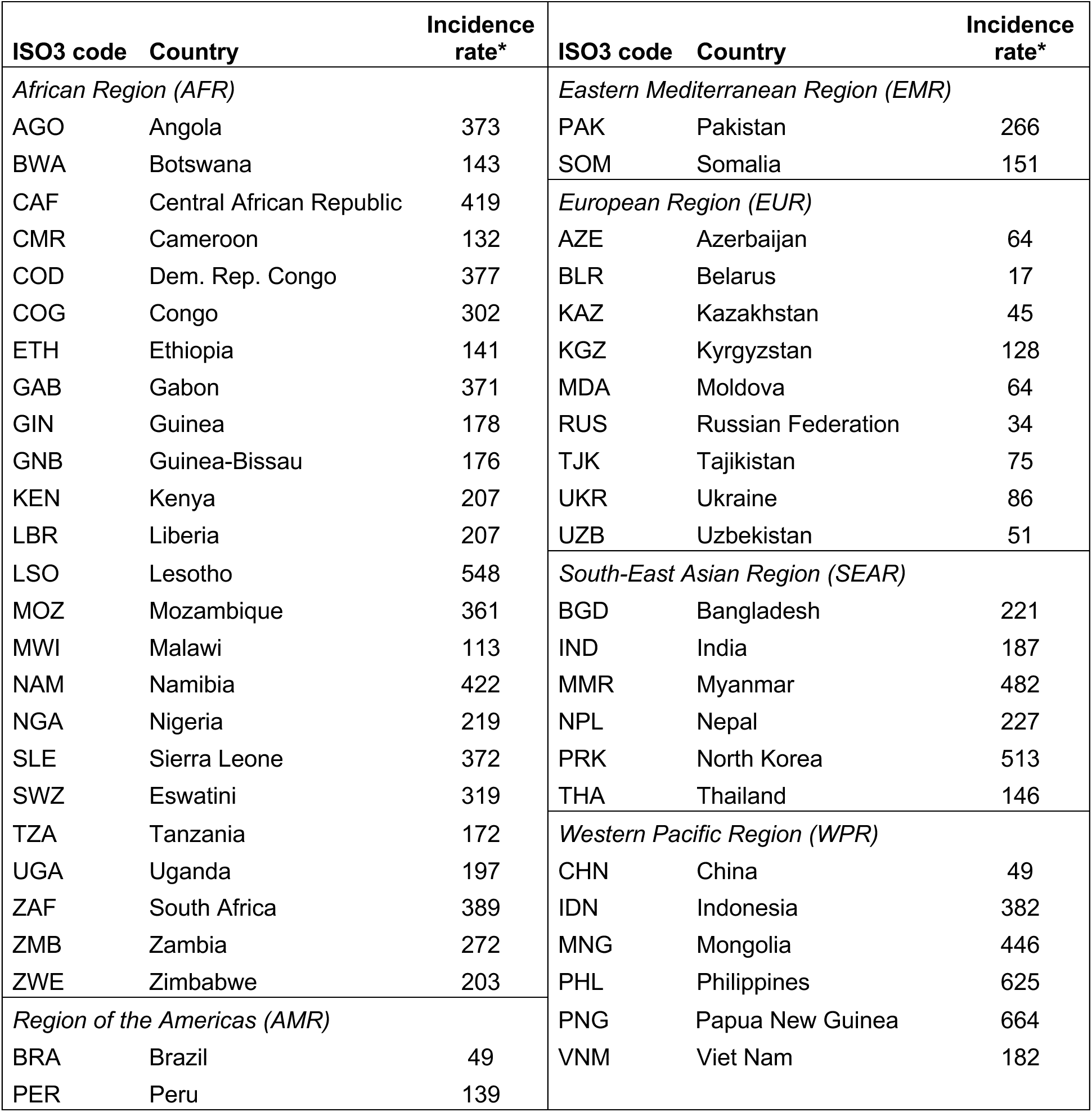
Origin countries included in the analysis. * TB incidence rate per 100,000, based on WHO epidemiological estimates for 2024.

| ISO3 code | Country | Incidence rate* | ISO3 code | Country | Incidence rate* |
| --- | --- | --- | --- | --- | --- |
| *African Region (AFR)* |  |  | *Eastern Mediterranean Region (EMR)* |  |  |
| AGO | Angola | 373 | PAK | Pakistan | 266 |
| BWA | Botswana | 143 | SOM | Somalia | 151 |
| CAF | Central African Republic | 419 | *European Region (EUR)* |  |  |
| CMR | Cameroon | 132 | AZE | Azerbaijan | 64 |
| COD | Dem. Rep. Congo | 377 | BLR | Belarus | 17 |
| COG | Congo | 302 | KAZ | Kazakhstan | 45 |
| ETH | Ethiopia | 141 | KGZ | Kyrgyzstan | 128 |
| GAB | Gabon | 371 | MDA | Moldova | 64 |
| GIN | Guinea | 178 | RUS | Russian Federation | 34 |
| GNB | Guinea-Bissau | 176 | TJK | Tajikistan | 75 |
| KEN | Kenya | 207 | UKR | Ukraine | 86 |
| LBR | Liberia | 207 | UZB | Uzbekistan | 51 |
| LSO | Lesotho | 548 | *South-East Asian Region (SEAR)* |  |  |
| MOZ | Mozambique | 361 | BGD | Bangladesh | 221 |
| MWI | Malawi | 113 | IND | India | 187 |
| NAM | Namibia | 422 | MMR | Myanmar | 482 |
| NGA | Nigeria | 219 | NPL | Nepal | 227 |
| SLE | Sierra Leone | 372 | PRK | North Korea | 513 |
| SWZ | Eswatini | 319 | THA | Thailand | 146 |
| TZA | Tanzania | 172 | *Western Pacific Region (WPR)* |  |  |
| UGA | Uganda | 197 | CHN | China | 49 |
| ZAF | South Africa | 389 | IDN | Indonesia | 382 |
| ZMB | Zambia | 272 | MNG | Mongolia | 446 |
| ZWE | Zimbabwe | 203 | PHL | Philippines | 625 |
| *Region of the Americas (AMR)* |  |  | PNG | Papua New Guinea | 664 |
| BRA | Brazil | 49 | VNM | Viet Nam | 182 |
| PER | Peru | 139 |  |  |  |

**Table S2:** Destination countries included in the analysis. * TB incidence rate per 100,000, based on WHO epidemiological estimates for 2024.

| ISO3 code | Country | Incidence rate* | ISO3 code | Country | Incidence rate* |
| --- | --- | --- | --- | --- | --- |
| *African Region (AFR)* |  |  | *European Region (EUR), continued* |  |  |
| SYC | Seychelles | 17 | GRC | Greece | 3.6 |
| *Region of the Americas (AMR)* |  |  | HRV | Croatia | 3.5 |
| ABW | Aruba | 8.3 | HUN | Hungary | 5.6 |
| ATG | Antigua and Barbuda | 1.1 | IRL | Ireland | 6.2 |
| BHS | Bahamas | 18 | ISL | Iceland | 3 |
| BRB | Barbados | 1 | ISR | Israel | 2.4 |
| CAN | Canada | 6.2 | ITA | Italy | 5 |
| CHL | Chile | 19 | LTU | Lithuania | 31 |
| CUW | Curaçao | 2.8 | LUX | Luxembourg | 7.5 |
| GUY | Guyana | 67 | LVA | Latvia | 21 |
| PAN | Panama | 63 | MLT | Malta | 15 |
| PRI | Puerto Rico | 0.55 | NLD | Netherlands | 4.7 |
| TTO | Trinidad and Tobago | 27 | NOR | Norway | 3.3 |
| URY | Uruguay | 47 | POL | Poland | 11 |
| USA | United States of America | 3.2 | PRT | Portugal | 15 |
| *Eastern Mediterranean Region (EMR)* |  |  | ROU | Romania | 55 |
| BHR | Bahrain | 12 | SVK | Slovakia | 3.3 |
| KWT | Kuwait | 10 | SVN | Slovenia | 2.6 |
| OMN | Oman | 11 | SWE | Sweden | 3.3 |
| QAT | Qatar | 37 | *South-East Asian Region (SEAR)* |  |  |
| SAU | Saudi Arabia | 8.4 | -- None -- |  |  |
| *European Region (EUR)* |  |  | *Western Pacific Region (WPR)* |  |  |
| AUT | Austria | 4.9 | AUS | Australia | 6.9 |
| BEL | Belgium | 2.7 | BRN | Brunei Darussalam | 65 |
| CHE | Switzerland | 5.4 | GUM | Guam | 55 |
| CYP | Cyprus | 5.5 | HKG | China, Hong Kong SAR | 66 |
| CZE | Czechia | 4.9 | JPN | Japan | 9.8 |
| DEU | Germany | 5.4 | KOR | Republic of Korea | 35 |
| DNK | Denmark | 3.2 | MAC | China, Macao SAR | 59 |
| ESP | Spain | 7.9 | NCL | New Caledonia | 7.9 |
| EST | Estonia | 8.2 | NZL | New Zealand | 24 |
| FIN | Finland | 3.7 | PYF | French Polynesia | 43 |
| FRA | France | 7.8 | SGP | Singapore | 6.9 |
| GBR | United Kingdom | 9.7 |  |  |  |

**Figure S1:**
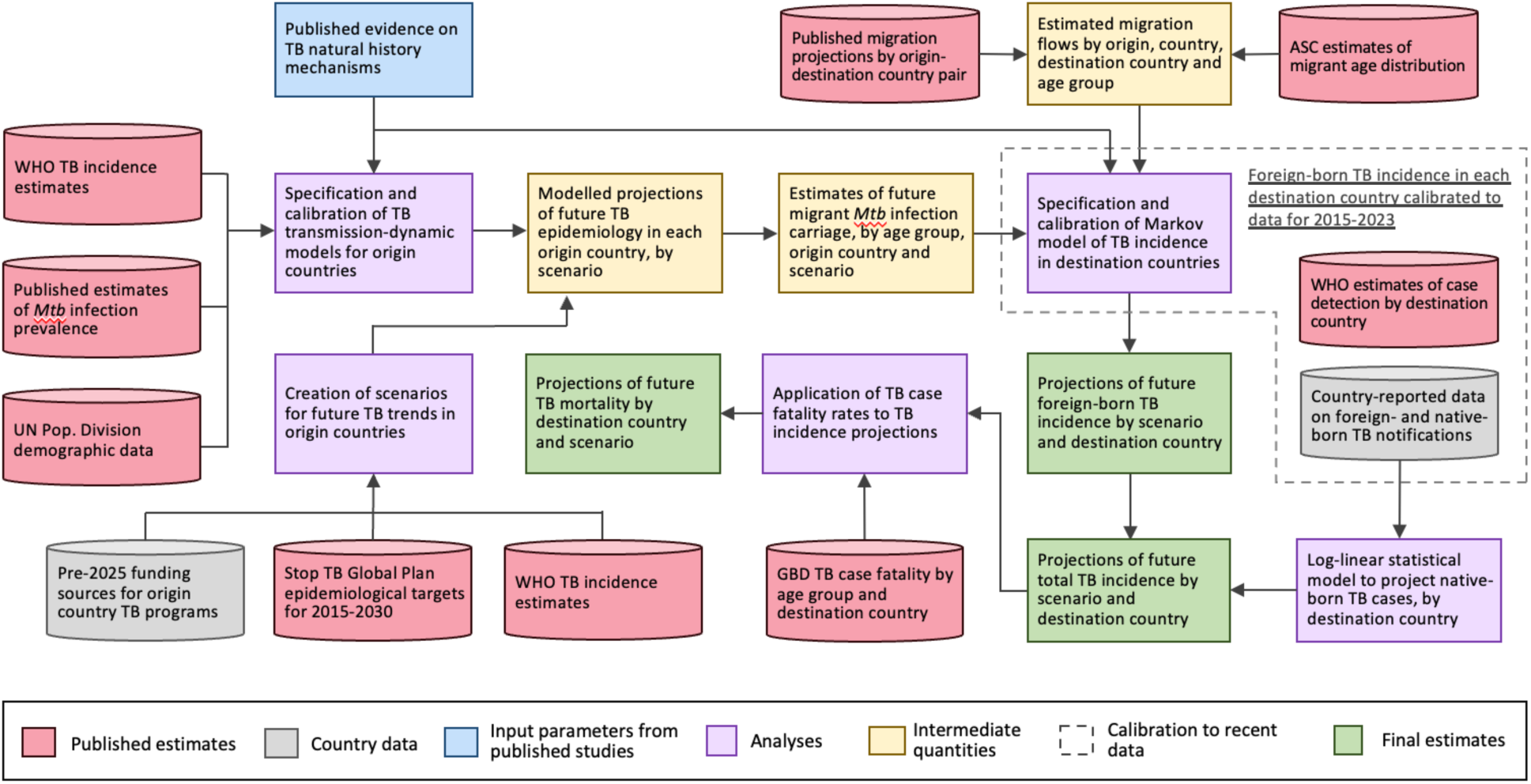
Schematic of data sources, analyses, and outcomes.

**Figure S2:**
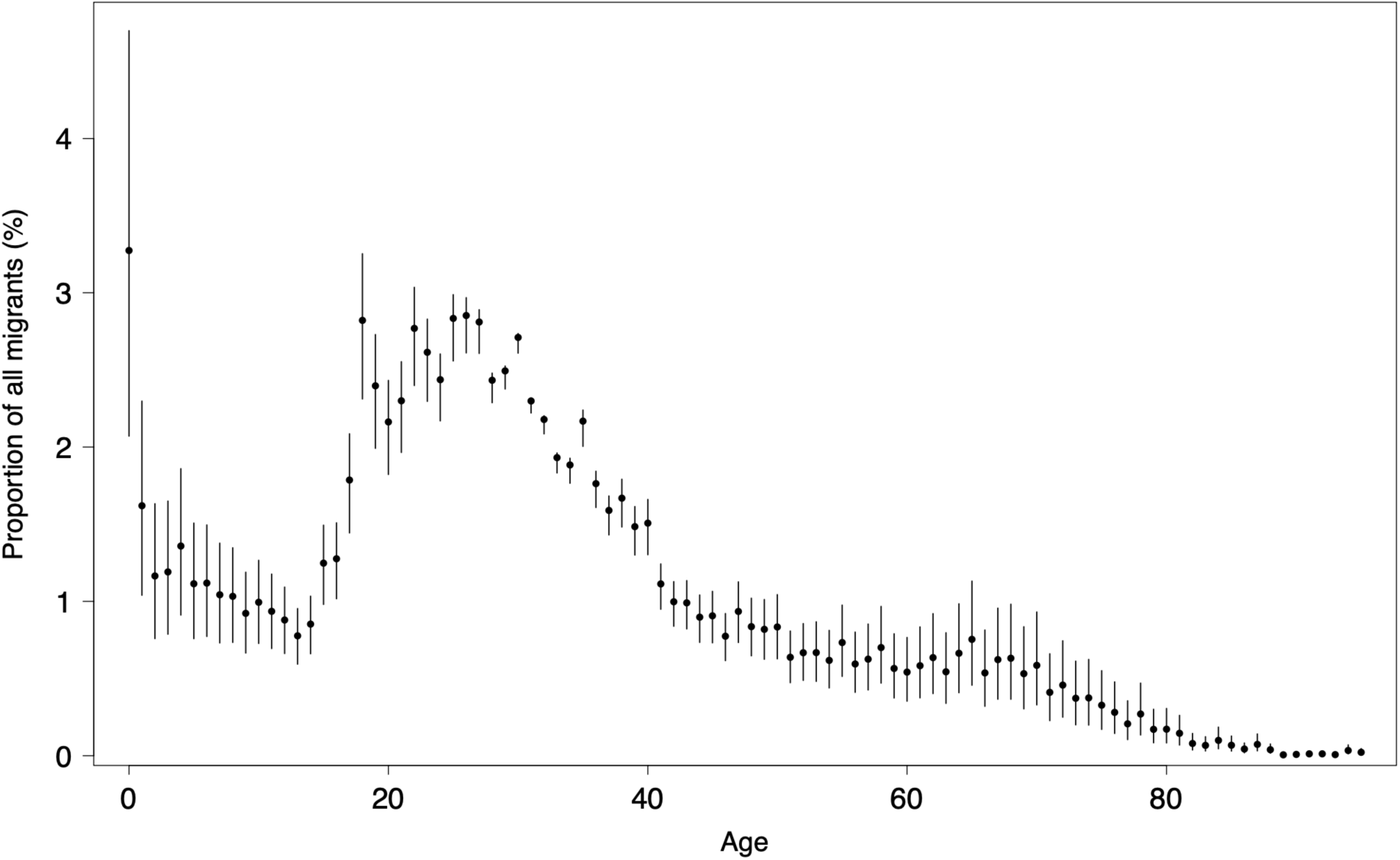
Age distribution assumed for migrant cohorts. Points represent mean values, bars represent 95% uncertainty intervals. Estimates based on American Community Survey data for new migrants, 2017-2021.

**Table S3:** Input data and sources.

| Input | Value (95% interval) | Prior distribution | Source |
| --- | --- | --- | --- |
| ***TB natural history*** |  |  |  |
| TB incidence rate for individuals with rapidly progressing *Mtb* infection | 0.55 (0.19, 1.10) | Gamma(5.44,9.89) | 33 |
| TB incidence rate for individuals with slowly progressing *Mtb* infection | 8.0 (4.0, 20.0) * 1e<sup>-4</sup> | Gamma(3.67, 4589) | 33 |
| Clearance rate for individuals with slowly progressing *Mtb* infection | 0.033 (0.020, 0.050) | Gamma(18.4, 558) | 33 |
| TB case fatality in destination countries | Stratified by country and age | Fixed values | 34 |
| ***Demography and migration*** |  |  |  |
| Origin country birth rates | Stratified by country and year | Fixed values | 30 |
| Origin country mortality rates | Stratified by country and age | Fixed values | 30 |
| Migration volume | Stratified by origin country, destination country, and year | Log Normal | 6 |
| Migrant mean age | 31 (26, 36) | Gamma(147,4.76) | 32 |
| Destination country mortality rates | Stratified by country and age | Fixed values | 30 |
| ***Native-born TB cases*** |  |  |  |
| Destination country native-born TB cases | Stratified by country and year | Log Normal | 7 |
| ***Calibration data*** |  |  |  |
| Origin country TB incidence trends | Stratified by country and year | --- | 27 |
| Origin country *Mtb* infection prevalence | Stratified by country | --- | 31 |
| Destination country foreign-born TB cases | Stratified by country and year | --- | 7 |
| ***Scenario specification*** |  |  |  |
| TB program funding sources for origin countries | Stratified by country and year | --- | 22 |
| Origin country TB incidence trends | Stratified by country and year | --- | 27 |

**Figure S3:**
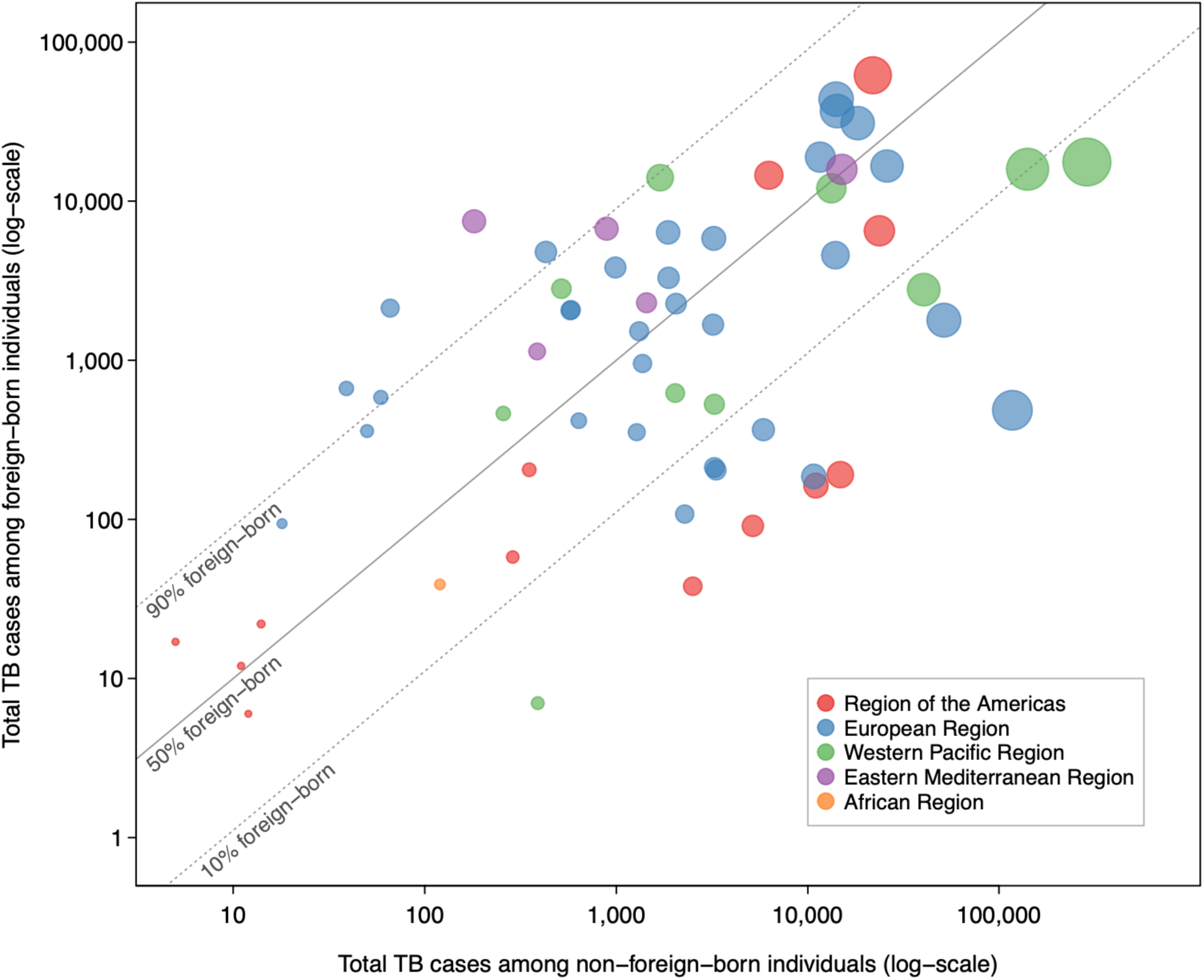
Scatterplot of total TB cases among foreign-born and non-foreign-born individuals over 2014-2024 in each HIC country included in the analysis.

**Figure S4:**
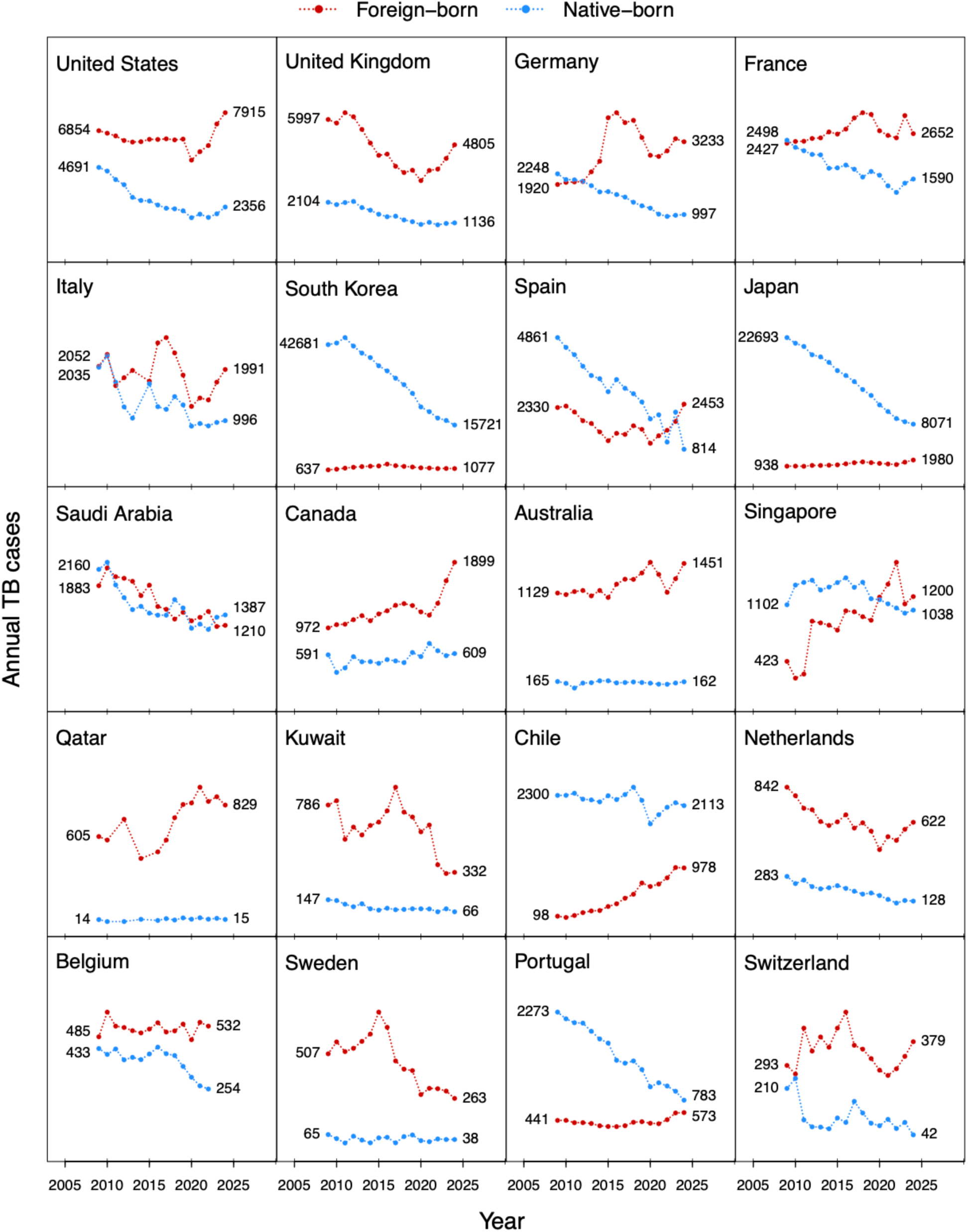
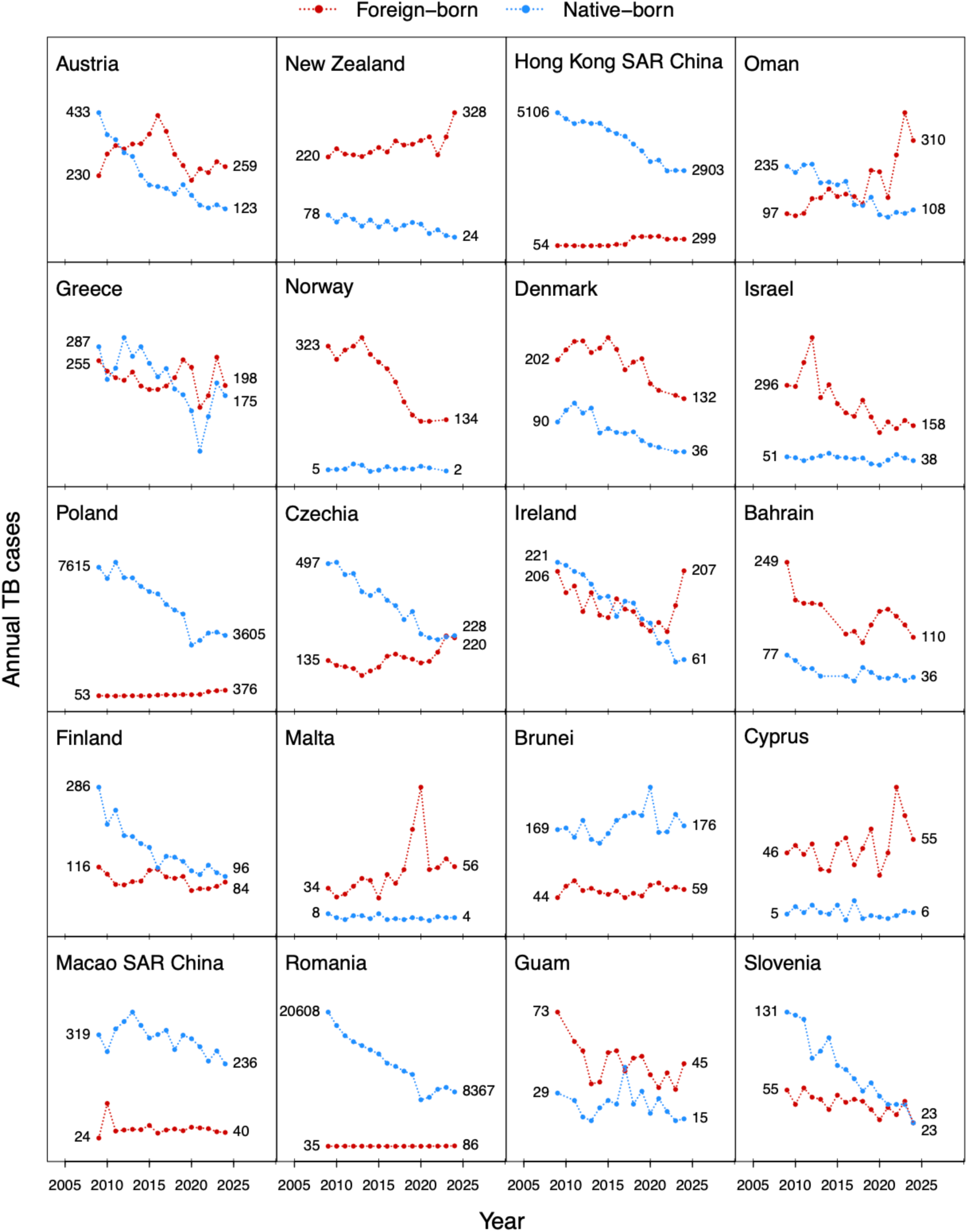
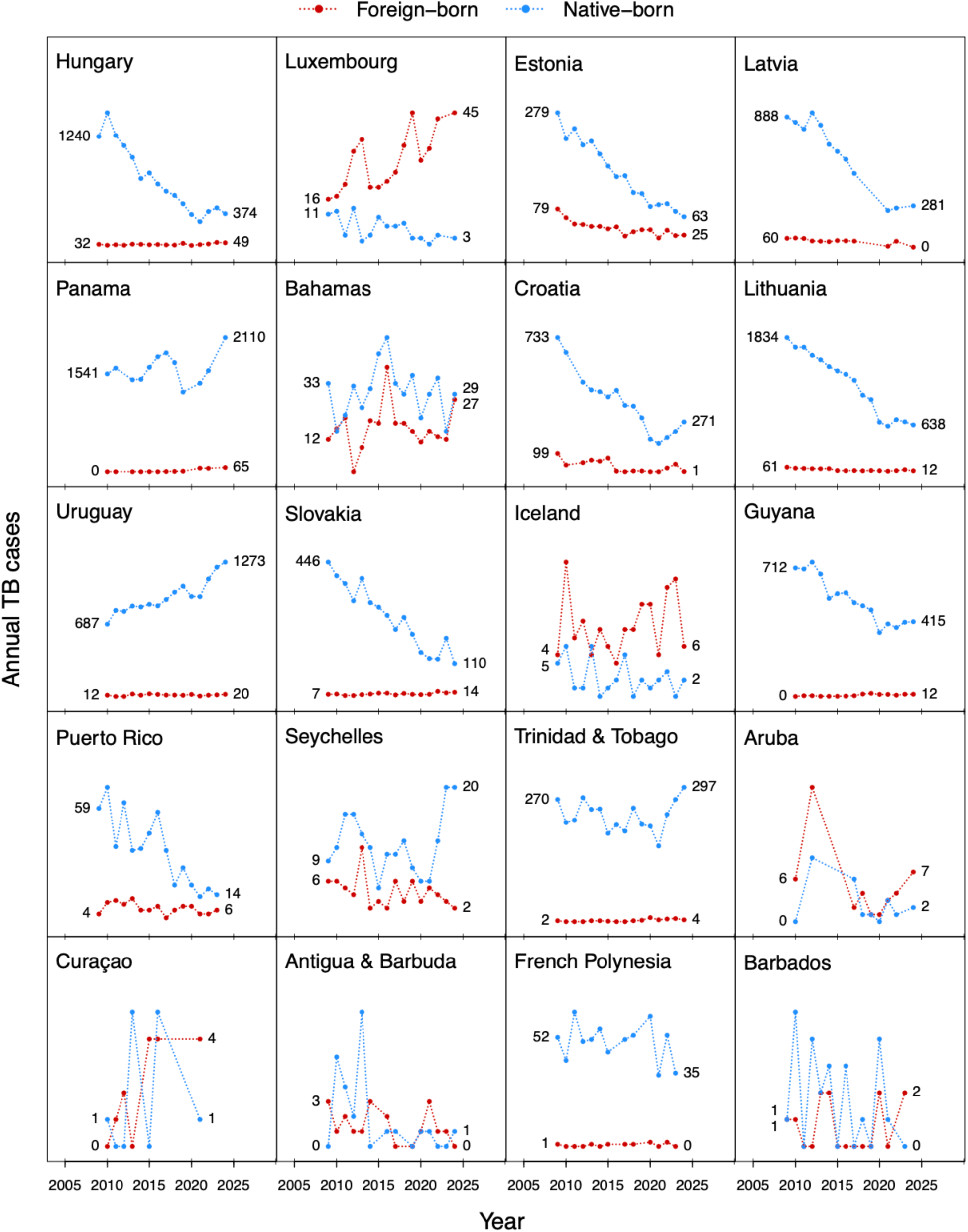
Time trends in TB among foreign-born and native-born individuals over 2009-2024 for each destination country.

**Figure S5:**
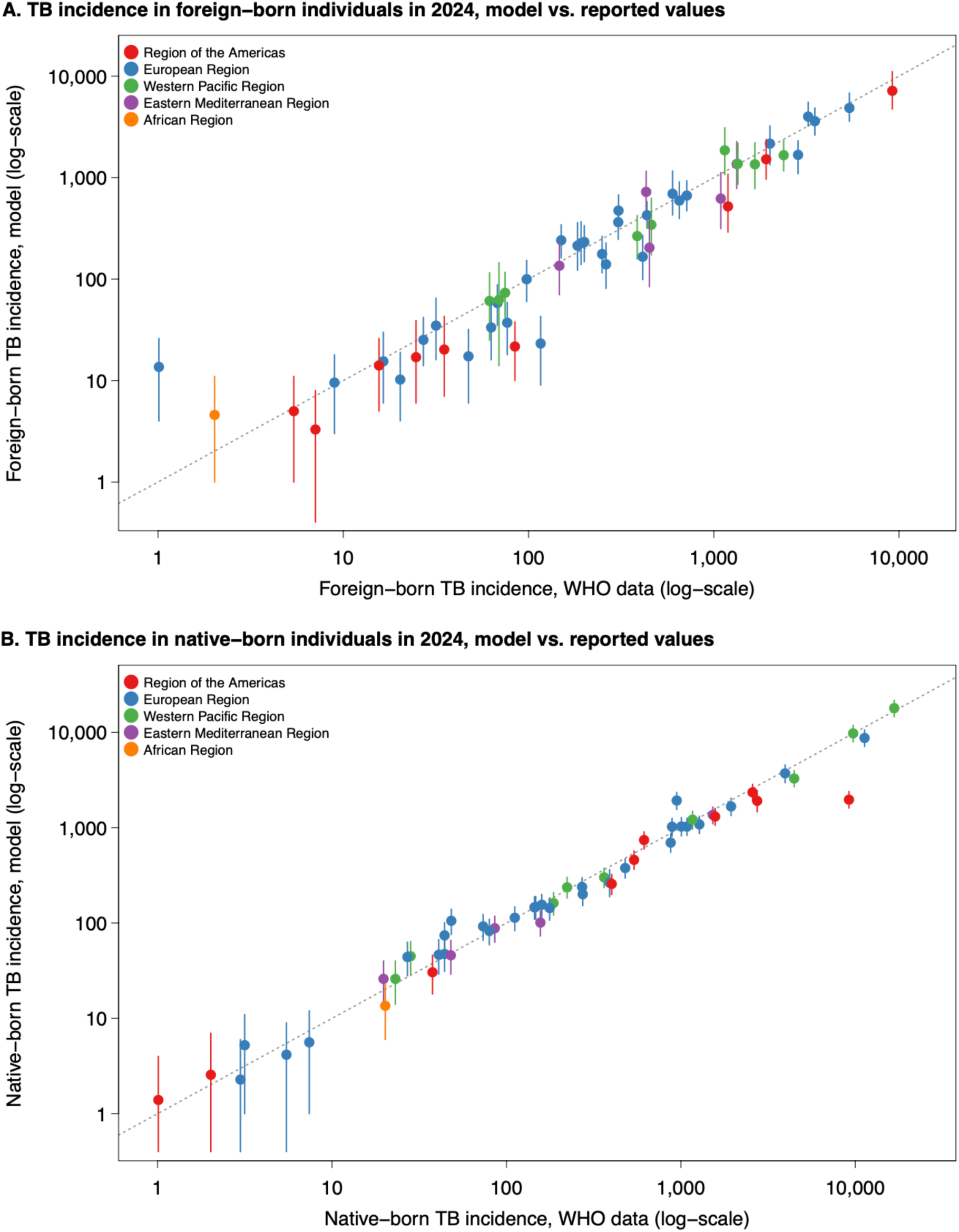
Validation of short-term model predictions, study estimates for 2024 compared to data reported to WHO for this year (not used for model fitting). Vertical lines represent 95% prediction intervals. Empirical incidence estimates calculated by dividing reported notifications (stratified by foreign-born vs. native-born) by WHO-estimated case detection rates for each country.

**Figure S6:**
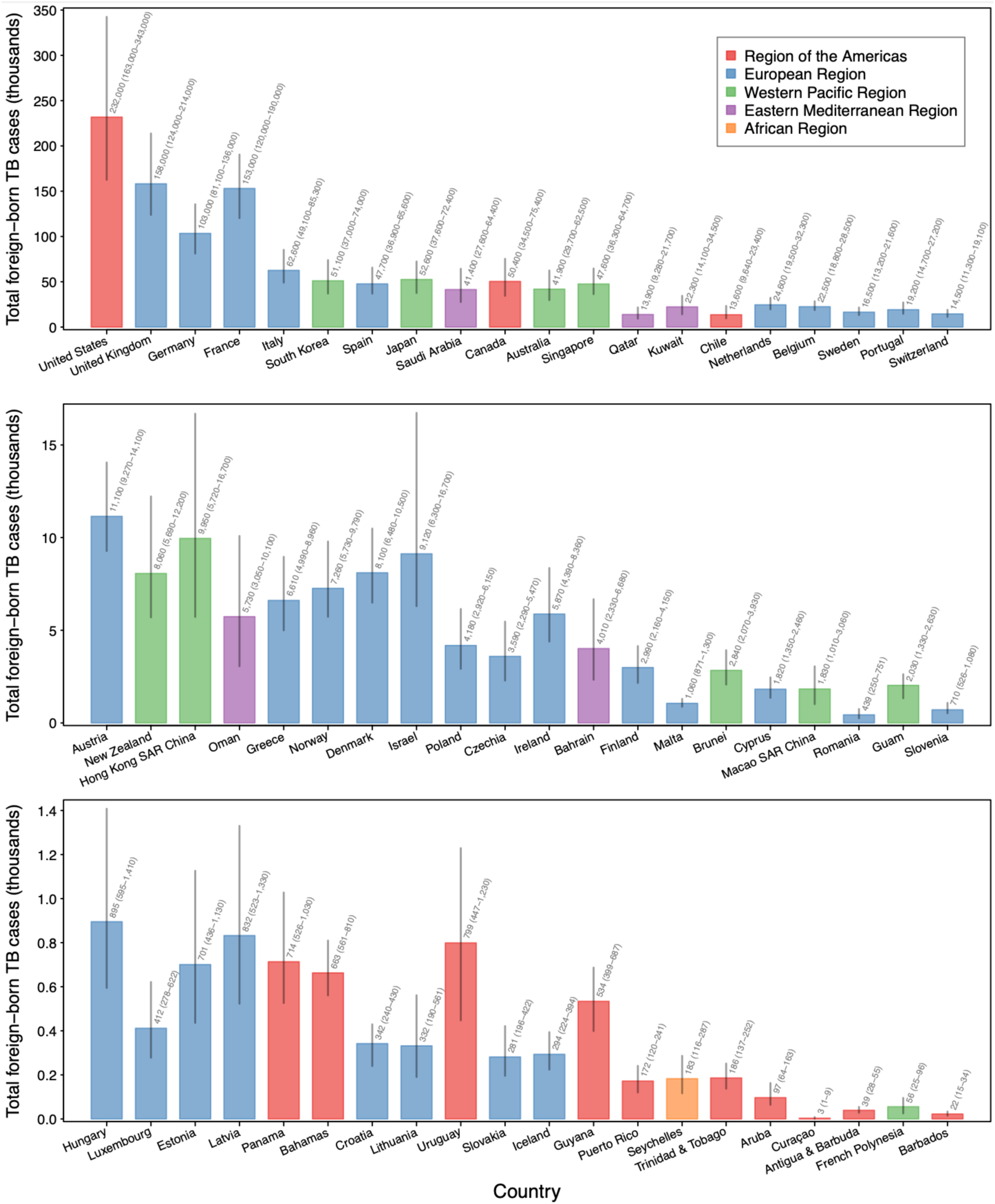
Projected numbers of foreign-born individuals developing TB over 2025-2050 for each destination country under the base-case scenario. Vertical lines represent 95% credible intervals. Countries ordered by total foreign-born TB cases reported for 2014-2024. Different y-axis scales used in each panel.

**Table S4:** Incremental numbers of TB cases among foreign-born individuals projected for 2025-2050 in each destination country under different scenarios, as compared to base-case. Countries ordered by total foreign-born TB notifications over 2014-2024. Positive values indicate additional TB cases as compared to the base-case. Negative values indicate fewer TB cases as compared to the base-case. Values in parentheses represent 95% uncertainty intervals.

| Country | Incremental number of TB cases, by scenario |  |  |  |
| --- | --- | --- | --- | --- |
|  | USAID and Global Fund support ended in 2025 | USAID support ended in 2025 | Cautious optimism | Achieve End TB Strategy targets |
| United States | 2.28 (1.80, 2.88) x1e5 | 1.06 (0.81, 1.29) x1e4 | -6.70 (-8.57, -5.31) x1e4 | -1.39 (-1.83, -1.06) x1e5 |
| United Kingdom | 1.54 (1.19, 1.84) x1e5 | 7.19 (5.85, 10.10) x1e3 | -4.64 (-5.34, -3.94) x1e4 | -9.64 (-11.76, -7.97) x1e4 |
| Germany | 1.51 (1.21, 1.89) x1e5 | 1.59 (1.29, 1.81) x1e4 | -3.00 (-3.55, -2.43) x1e4 | -6.20 (-7.47, -5.01) x1e4 |
| France | 1.68 (1.19, 2.18) x1e5 | 1.98 (1.39, 2.69) x1e4 | -4.39 (-5.21, -3.47) x1e4 | -9.98 (-12.09, -7.71) x1e4 |
| Italy | 5.06 (3.94, 6.23) x1e4 | 3.34 (2.85, 3.79) x1e3 | -1.78 (-2.02, -1.54) x1e4 | -3.60 (-4.29, -3.02) x1e4 |
| South Korea | 2.29 (1.55, 2.91) x1e4 | 3.65 (2.58, 5.42) x1e3 | -1.51 (-2.25, -1.09) x1e4 | -3.22 (-5.05, -2.28) x1e4 |
| Spain | 3.26 (2.72, 3.90) x1e4 | 3.81 (2.60, 5.73) x1e3 | -1.35 (-1.76, -1.10) x1e4 | -2.66 (-3.46, -2.14) x1e4 |
| Japan | 2.06 (1.57, 2.57) x1e4 | 2.63 (1.90, 3.59) x1e3 | -1.57 (-1.98, -1.14) x1e4 | -3.09 (-4.00, -2.22) x1e4 |
| Saudi Arabia | 2.19 (1.78, 2.68) x1e4 | 3.89 (2.72, 4.80) x1e3 | -1.06 (-1.28, -0.87) x1e4 | -2.39 (-3.22, -1.83) x1e4 |
| Canada | 4.07 (3.30, 4.81) x1e4 | 2.30 (1.83, 2.86) x1e3 | -1.45 (-1.77, -1.15) x1e4 | -3.11 (-4.19, -2.30) x1e4 |
| Australia | 2.70 (2.23, 3.40) x1e4 | 2.09 (1.61, 2.70) x1e3 | -1.19 (-1.53, -0.97) x1e4 | -2.52 (-3.45, -1.94) x1e4 |
| Singapore | 1.79 (1.31, 2.34) x1e4 | 4.05 (2.75, 4.97) x1e3 | -1.44 (-1.81, -1.16) x1e4 | -3.15 (-4.06, -2.51) x1e4 |
| Qatar | 4.49 (3.49, 5.91) x1e3 | 7.68 (5.61, 10.87) x1e2 | -3.45 (-4.29, -2.50) x1e3 | -7.31 (-9.51, -5.16) x1e3 |
| Kuwait | 9.86 (7.39, 14.03) x1e3 | 8.33 (6.40, 10.18) x1e2 | -6.26 (-11.04, -3.92) x1e3 | -1.34 (-2.26, -0.79) x1e4 |
| Chile | 9.65 (5.44, 18.81) x1e3 | 2.75 (1.49, 5.33) x1e3 | -3.94 (-7.02, -2.73) x1e3 | -7.90 (-14.44, -5.27) x1e3 |
| Netherlands | 2.25 (1.79, 2.86) x1e4 | 3.03 (2.18, 3.69) x1e3 | -7.44 (-8.69, -5.87) x1e3 | -1.60 (-1.97, -1.20) x1e4 |
| Belgium | 4.88 (3.38, 7.41) x1e4 | 4.21 (2.90, 6.21) x1e3 | -6.66 (-7.89, -5.58) x1e3 | -1.43 (-1.74, -1.18) x1e4 |
| Sweden | 1.81 (1.51, 2.12) x1e4 | 1.02 (0.83, 1.20) x1e3 | -4.87 (-6.03, -4.12) x1e3 | -1.02 (-1.26, -0.84) x1e4 |
| Portugal | 7.16 (5.86, 8.63) x1e3 | 3.34 (2.50, 4.24) x1e2 | -5.60 (-7.46, -4.53) x1e3 | -1.05 (-1.40, -0.84) x1e4 |
| Switzerland | 1.44 (1.09, 2.35) x1e4 | 1.00 (0.72, 1.44) x1e3 | -4.27 (-5.12, -3.52) x1e3 | -8.81 (-11.08, -7.03) x1e3 |
| Austria | 1.11 (0.93, 1.29) x1e4 | 7.24 (5.53, 9.35) x1e2 | -3.24 (-3.57, -2.80) x1e3 | -6.60 (-7.53, -5.68) x1e3 |
| New Zealand | 3.44 (2.66, 4.12) x1e3 | 3.40 (2.74, 4.28) x1e2 | -2.27 (-2.70, -1.88) x1e3 | -4.92 (-6.44, -3.85) x1e3 |
| Hong Kong SAR China | 2.03 (1.45, 2.63) x1e3 | 7.09 (4.92, 9.27) x1e2 | -2.44 (-3.18, -1.69) x1e3 | -5.69 (-8.53, -3.57) x1e3 |
| Oman | 1.52 (1.13, 2.18) x1e3 | 9.98 (6.40, 19.09) x1e1 | -1.39 (-1.96, -0.91) x1e3 | -3.12 (-4.82, -1.89) x1e3 |
| Greece | 9.69 (7.76, 11.46) x1e3 | 8.45 (6.71, 10.26) x1e2 | -1.91 (-2.26, -1.51) x1e3 | -4.01 (-4.89, -3.09) x1e3 |
| Norway | 8.67 (7.42, 10.03) x1e3 | 4.25 (3.55, 4.95) x1e2 | -2.10 (-2.38, -1.80) x1e3 | -4.39 (-5.30, -3.66) x1e3 |
| Denmark | 7.95 (6.77, 10.09) x1e3 | 4.63 (3.89, 5.59) x1e2 | -2.43 (-2.79, -2.05) x1e3 | -5.08 (-6.00, -4.22) x1e3 |
| Israel | 1.32 (1.03, 2.14) x1e4 | 1.04 (0.82, 1.73) x1e3 | -2.71 (-4.59, -2.10) x1e3 | -5.87 (-9.60, -4.25) x1e3 |
| Poland | 4.99 (4.18, 6.15) x1e3 | 3.70 (3.01, 4.64) x1e2 | -1.09 (-1.27, -0.93) x1e3 | -2.06 (-2.57, -1.71) x1e3 |
| Czechia | 7.05 (4.44, 10.18) x1e3 | 2.12 (1.69, 2.81) x1e2 | -9.12 (-11.88, -6.73) x1e2 | -1.70 (-2.27, -1.24) x1e3 |

**Table S4 continued: Incremental numbers of TB cases among foreign-born individuals projected for 2025-2050 in each destination country under different scenarios, as compared to base-case.**
| Country | Incremental number of TB cases, by scenario |  |  |  |
| --- | --- | --- | --- | --- |
|  | USAID and Global Fund support ended in 2025 | USAID support ended in 2025 | Cautious optimism | Achieve End TB Strategy targets |
| Ireland | 7.67 (6.02, 9.85) x1e3 | 1.72 (1.34, 2.14) x1e2 | -1.72 (-2.12, -1.41) x1e3 | -3.35 (-4.26, -2.70) x1e3 |
| Bahrain | 1.42 (1.10, 1.86) x1e3 | 1.09 (0.72, 1.61) x1e2 | -1.03 (-1.41, -0.74) x1e3 | -2.29 (-3.54, -1.49) x1e3 |
| Finland | 4.19 (3.51, 4.86) x1e3 | 2.08 (1.45, 2.63) x1e2 | -8.50 (-10.18, -6.56) x1e2 | -1.75 (-2.21, -1.30) x1e3 |
| Malta | 6.61 (5.28, 7.67) x1e2 | 5.73 (4.38, 7.14) x1e1 | -2.84 (-3.27, -2.35) x1e2 | -6.21 (-7.41, -4.99) x1e2 |
| Brunei | 8.02 (5.02, 12.72) x1e2 | 2.24 (1.43, 3.42) x1e2 | -9.01 (-12.15, -6.93) x1e2 | -1.98 (-2.72, -1.49) x1e3 |
| Cyprus | 1.75 (1.29, 2.08) x1e3 | 1.77 (1.28, 2.14) x1e2 | -5.55 (-6.77, -4.34) x1e2 | -1.17 (-1.48, -0.90) x1e3 |
| Macao SAR China | 2.79 (1.64, 4.78) x1e2 | 1.12 (0.66, 1.90) x1e2 | -4.83 (-8.20, -2.84) x1e2 | -1.09 (-1.94, -0.62) x1e3 |
| Romania | 5.04 (3.71, 6.95) x1e2 | 1.76 (1.33, 2.52) x1e1 | -8.19 (-10.44, -6.24) x1e1 | -1.75 (-2.42, -1.23) x1e2 |
| Guam | 1.32 (1.01, 1.61) x1e2 | 7.15 (4.68, 9.75) x1e1 | -6.41 (-8.95, -4.36) x1e2 | -1.43 (-2.03, -0.94) x1e3 |
| Slovenia | 3.26 (2.55, 3.98) x1e2 | 6.83 (5.06, 9.54) x1e1 | -1.86 (-3.28, -1.35) x1e2 | -4.23 (-8.11, -2.89) x1e2 |
| Hungary | 1.18 (0.98, 1.42) x1e3 | 5.61 (4.60, 7.30) x1e1 | -2.27 (-2.81, -1.83) x1e2 | -4.55 (-6.08, -3.50) x1e2 |
| Luxembourg | 1.99 (1.51, 2.78) x1e2 | 1.37 (0.88, 1.86) x1e1 | -1.05 (-1.60, -0.82) x1e2 | -2.05 (-3.04, -1.56) x1e2 |
| Estonia | 2.83 (1.99, 3.39) x1e3 | 1.58 (1.14, 1.90) x1e2 | -1.56 (-1.89, -1.16) x1e2 | -3.36 (-4.48, -2.40) x1e2 |
| Latvia | 3.15 (2.62, 3.79) x1e3 | 1.71 (1.41, 2.05) x1e2 | -1.89 (-2.43, -1.55) x1e2 | -4.03 (-5.67, -3.08) x1e2 |
| Panama | 2.13 (1.67, 2.61) x1e2 | 5.03 (3.86, 6.27) x1e1 | -2.03 (-2.39, -1.70) x1e2 | -4.28 (-5.49, -3.42) x1e2 |
| Bahamas | 2.20 (1.79, 2.78) x1e2 | 2.63 (1.47, 5.06) x1e1 | -1.80 (-2.22, -1.48) x1e2 | -4.05 (-5.35, -3.20) x1e2 |
| Croatia | 1.86 (1.17, 2.71) x1e2 | 3.09 (2.13, 4.45) x1e1 | -8.85 (-10.75, -6.05) x1e1 | -1.89 (-2.32, -1.24) x1e2 |
| Lithuania | 1.23 (0.92, 1.55) x1e3 | 6.20 (4.58, 7.82) x1e1 | -7.05 (-9.00, -5.35) x1e1 | -1.54 (-2.21, -1.06) x1e2 |
| Uruguay | 1.55 (0.94, 2.33) x1e2 | 0 (0, 0) | -2.59 (-3.89, -1.55) x1e2 | -4.42 (-6.88, -2.56) x1e2 |
| Slovakia | 4.39 (3.03, 6.13) x1e2 | 1.76 (1.46, 2.01) x1e1 | -7.48 (-9.37, -5.74) x1e1 | -1.43 (-1.90, -1.08) x1e2 |
| Iceland | 1.89 (1.59, 2.39) x1e2 | 1.48 (1.19, 1.89) x1e1 | -8.88 (-11.75, -6.92) x1e1 | -1.82 (-2.43, -1.37) x1e2 |
| Guyana | 6.03 (4.69, 7.79) x1e1 | 6.11 (4.80, 7.55) x1e0 | -1.39 (-1.85, -1.06) x1e2 | -2.64 (-3.50, -2.01) x1e2 |
| Puerto Rico | 8.56 (5.84, 12.02) x1e1 | 2.59 (1.82, 3.49) x1e1 | -5.27 (-6.20, -3.67) x1e1 | -1.11 (-1.39, -0.73) x1e2 |
| Seychelles | 5.30 (3.85, 7.23) x1e1 | 1.52 (0.97, 1.98) x1e0 | -5.03 (-6.97, -3.25) x1e1 | -9.58 (-13.94, -5.99) x1e1 |
| Trinidad & Tobago | 1.57 (1.08, 2.28) x1e2 | 3.11 (1.70, 4.13) x1e0 | -5.54 (-7.07, -4.12) x1e1 | -1.11 (-1.41, -0.82) x1e2 |
| Aruba | 4.36 (2.89, 6.98) x1e1 | 1.25 (0.83, 2.12) x1e1 | -2.98 (-5.59, -2.09) x1e1 | -6.41 (-12.27, -4.29) x1e1 |
| Antigua & Barbuda | 3.22 (2.39, 4.18) x1e1 | 0 (0, 0) | -1.11 (-1.37, -0.79) x1e1 | -2.14 (-2.70, -1.49) x1e1 |
| French Polynesia | 8.72 (3.46, 17.19) x1e0 | 3.58 (1.44, 6.98) x1e0 | -1.80 (-3.55, -0.72) x1e1 | -4.07 (-8.01, -1.52) x1e1 |
| Barbados | 3.37 (2.38, 4.86) x1e0 | 0 (0, 0) | -6.78 (-9.71, -4.85) x1e0 | -1.47 (-2.08, -0.98) x1e1 |

**Table S5:** Incremental numbers of TB deaths among foreign-born individuals projected for 2025-2050 in each destination country under different scenarios, as compared to base-case. Countries ordered by total foreign-born TB notifications over 2014-2024. Positive values indicate additional TB deaths as compared to the base-case. Negative values indicate fewer TB cases as compared to the base-case. Values in parentheses represent 95% uncertainty intervals.

| Country | Incremental number of TB deaths, by scenario |  |  |  |
| --- | --- | --- | --- | --- |
|  | USAID and Global Fund support ended in 2025 | USAID support ended in 2025 | Cautious optimism | Achieve End TB Strategy targets |
| United States | 1.08 (0.87, 1.49) x1e4 | 4.99 (4.04, 5.84) x1e2 | -3.15 (-3.79, -2.62) x1e3 | -6.92 (-9.23, -5.28) x1e3 |
| United Kingdom | 1.46 (1.19, 1.75) x1e4 | 6.67 (5.39, 8.71) x1e2 | -4.32 (-5.05, -3.62) x1e3 | -9.26 (-12.03, -7.26) x1e3 |
| Germany | 1.43 (1.20, 1.69) x1e4 | 1.48 (1.23, 1.68) x1e3 | -2.77 (-3.11, -2.35) x1e3 | -5.87 (-7.00, -4.87) x1e3 |
| France | 1.61 (1.12, 2.05) x1e4 | 1.85 (1.28, 2.52) x1e3 | -4.06 (-4.81, -3.18) x1e3 | -9.26 (-11.42, -7.26) x1e3 |
| Italy | 4.79 (3.97, 5.75) x1e3 | 3.14 (2.63, 3.56) x1e2 | -1.66 (-1.87, -1.40) x1e3 | -3.45 (-4.30, -2.79) x1e3 |
| South Korea | 1.41 (0.94, 1.79) x1e3 | 2.18 (1.52, 3.22) x1e2 | -8.92 (-13.12, -6.46) x1e2 | -1.94 (-3.05, -1.35) x1e3 |
| Spain | 3.08 (2.52, 3.98) x1e3 | 3.58 (2.42, 5.75) x1e2 | -1.25 (-1.57, -0.99) x1e3 | -2.51 (-3.17, -1.90) x1e3 |
| Japan | 1.25 (0.97, 1.56) x1e3 | 1.56 (1.15, 2.08) x1e2 | -9.31 (-11.67, -6.92) x1e2 | -1.87 (-2.39, -1.38) x1e3 |
| Saudi Arabia | 2.03 (1.59, 2.55) x1e3 | 3.60 (2.32, 4.48) x1e2 | -9.85 (-11.91, -7.86) x1e2 | -2.32 (-3.29, -1.67) x1e3 |
| Canada | 1.94 (1.59, 2.42) x1e3 | 1.10 (0.88, 1.38) x1e2 | -6.88 (-8.36, -5.20) x1e2 | -1.58 (-2.23, -1.08) x1e3 |
| Australia | 1.63 (1.29, 2.07) x1e3 | 1.24 (0.96, 1.57) x1e2 | -7.00 (-8.76, -5.47) x1e2 | -1.54 (-2.14, -1.09) x1e3 |
| Singapore | 1.08 (0.79, 1.42) x1e3 | 2.43 (1.68, 2.98) x1e2 | -8.60 (-10.68, -6.90) x1e2 | -1.90 (-2.43, -1.49) x1e3 |
| Qatar | 4.11 (3.17, 5.24) x1e2 | 6.93 (5.07, 9.51) x1e1 | -3.15 (-3.89, -2.26) x1e2 | -6.90 (-9.49, -4.70) x1e2 |
| Kuwait | 9.07 (6.58, 12.96) x1e2 | 7.64 (5.51, 9.20) x1e1 | -5.75 (-10.14, -3.34) x1e2 | -1.28 (-2.14, -0.68) x1e3 |
| Chile | 1.43 (0.85, 2.70) x1e3 | 4.05 (2.30, 7.59) x1e2 | -5.82 (-9.99, -4.03) x1e2 | -1.19 (-2.12, -0.80) x1e3 |
| Netherlands | 2.13 (1.66, 2.76) x1e3 | 2.81 (1.94, 3.51) x1e2 | -6.89 (-7.90, -5.40) x1e2 | -1.52 (-1.95, -1.12) x1e3 |
| Belgium | 4.68 (3.06, 6.78) x1e3 | 3.99 (2.60, 5.60) x1e2 | -6.21 (-7.09, -5.16) x1e2 | -1.36 (-1.64, -1.10) x1e3 |
| Sweden | 1.72 (1.51, 1.92) x1e3 | 9.49 (7.69, 10.65) x1e1 | -4.52 (-5.35, -3.89) x1e2 | -9.64 (-11.81, -8.02) x1e2 |
| Portugal | 6.80 (5.33, 8.24) x1e2 | 3.16 (2.48, 3.97) x1e1 | -5.23 (-6.72, -4.24) x1e2 | -1.01 (-1.35, -0.77) x1e3 |
| Switzerland | 1.37 (1.04, 2.09) x1e3 | 9.36 (6.97, 12.95) x1e1 | -3.96 (-4.57, -3.33) x1e2 | -8.38 (-10.38, -6.72) x1e2 |
| Austria | 1.05 (0.91, 1.18) x1e3 | 6.69 (5.31, 8.50) x1e1 | -2.99 (-3.27, -2.60) x1e2 | -6.20 (-7.27, -5.24) x1e2 |
| New Zealand | 2.08 (1.61, 2.63) x1e2 | 2.02 (1.59, 2.71) x1e1 | -1.34 (-1.60, -1.07) x1e2 | -3.00 (-4.09, -2.19) x1e2 |
| Hong Kong SAR China | 1.21 (0.83, 1.60) x1e2 | 4.23 (2.82, 5.60) x1e1 | -1.46 (-1.98, -0.95) x1e2 | -3.66 (-5.93, -2.16) x1e2 |
| Oman | 1.42 (1.01, 2.16) x1e2 | 9.28 (6.37, 17.04) x1e0 | -1.30 (-1.88, -0.86) x1e2 | -3.10 (-5.10, -1.75) x1e2 |
| Greece | 9.16 (7.62, 10.79) x1e2 | 7.84 (6.50, 9.16) x1e1 | -1.77 (-2.07, -1.48) x1e2 | -3.83 (-4.83, -3.04) x1e2 |
| Norway | 8.25 (7.03, 10.07) x1e2 | 3.98 (3.33, 4.77) x1e1 | -1.96 (-2.33, -1.68) x1e2 | -4.22 (-5.37, -3.39) x1e2 |
| Denmark | 7.56 (6.16, 10.23) x1e2 | 4.31 (3.54, 5.49) x1e1 | -2.25 (-2.75, -1.88) x1e2 | -4.82 (-6.01, -3.92) x1e2 |
| Israel | 1.24 (0.91, 1.98) x1e3 | 9.55 (7.14, 15.77) x1e1 | -2.48 (-4.15, -1.79) x1e2 | -5.51 (-8.89, -3.66) x1e2 |
| Poland | 4.73 (3.85, 6.16) x1e2 | 3.46 (2.77, 4.57) x1e1 | -1.02 (-1.23, -0.85) x1e2 | -1.99 (-2.62, -1.55) x1e2 |
| Czechia | 6.66 (4.38, 10.34) x1e2 | 1.97 (1.59, 2.50) x1e1 | -8.52 (-11.48, -6.53) x1e1 | -1.64 (-2.29, -1.22) x1e2 |

**Table S5 continued: Incremental numbers of TB deaths among foreign-born individuals projected for 2025-2050 in each destination country under different scenarios, as compared to base-case.**
| Country | Incremental number of TB deaths, by scenario |  |  |  |
| --- | --- | --- | --- | --- |
|  | USAID and Global Fund support ended in 2025 | USAID support ended in 2025 | Cautious optimism | Achieve End TB Strategy targets |
| Ireland | 7.27 (5.86, 9.02) x1e2 | 1.61 (1.32, 2.07) x1e1 | -1.60 (-1.93, -1.34) x1e2 | -3.22 (-4.12, -2.59) x1e2 |
| Bahrain | 1.32 (0.96, 1.78) x1e2 | 1.01 (0.71, 1.53) x1e1 | -9.64 (-13.63, -6.50) x1e1 | -2.24 (-3.67, -1.34) x1e2 |
| Finland | 3.98 (3.46, 4.58) x1e2 | 1.95 (1.46, 2.37) x1e1 | -7.94 (-9.49, -6.56) x1e1 | -1.69 (-2.19, -1.33) x1e2 |
| Malta | 6.17 (4.87, 7.47) x1e1 | 5.19 (3.92, 6.36) x1e0 | -2.58 (-2.92, -2.11) x1e1 | -5.63 (-6.62, -4.47) x1e1 |
| Brunei | 4.84 (3.06, 7.61) x1e1 | 1.32 (0.86, 2.03) x1e1 | -5.35 (-7.22, -4.07) x1e1 | -1.18 (-1.61, -0.88) x1e2 |
| Cyprus | 1.65 (1.31, 1.90) x1e2 | 1.64 (1.28, 2.05) x1e1 | -5.12 (-5.99, -4.18) x1e1 | -1.11 (-1.37, -0.87) x1e2 |
| Macao SAR China | 1.66 (1.00, 2.65) x1e1 | 6.64 (4.04, 10.53) x1e0 | -2.87 (-4.55, -1.75) x1e1 | -6.88 (-12.16, -3.84) x1e1 |
| Romania | 4.84 (3.33, 6.60) x1e1 | 1.69 (1.23, 2.52) x1e0 | -7.90 (-10.41, -5.53) x1e0 | -1.78 (-2.68, -1.12) x1e1 |
| Guam | 7.94 (6.16, 9.75) x1e0 | 4.19 (2.80, 5.67) x1e0 | -3.86 (-5.44, -2.63) x1e1 | -8.56 (-12.22, -5.68) x1e1 |
| Slovenia | 3.08 (2.56, 3.78) x1e1 | 6.27 (4.94, 8.86) x1e0 | -1.70 (-3.04, -1.27) x1e1 | -3.86 (-7.45, -2.71) x1e1 |
| Hungary | 1.12 (0.94, 1.30) x1e2 | 5.30 (4.39, 6.49) x1e0 | -2.15 (-2.68, -1.78) x1e1 | -4.53 (-6.39, -3.41) x1e1 |
| Luxembourg | 1.89 (1.40, 2.47) x1e1 | 1.29 (0.83, 1.65) x1e0 | -9.89 (-15.73, -7.25) x1e0 | -1.98 (-3.08, -1.40) x1e1 |
| Estonia | 2.69 (2.06, 3.19) x1e2 | 1.50 (1.18, 1.77) x1e1 | -1.49 (-1.82, -1.19) x1e1 | -3.42 (-4.83, -2.52) x1e1 |
| Latvia | 3.02 (2.41, 3.72) x1e2 | 1.64 (1.31, 2.02) x1e1 | -1.81 (-2.47, -1.45) x1e1 | -4.13 (-6.28, -2.91) x1e1 |
| Panama | 3.25 (2.47, 3.97) x1e1 | 7.70 (5.64, 9.77) x1e0 | -3.09 (-3.73, -2.51) x1e1 | -6.83 (-9.41, -5.04) x1e1 |
| Bahamas | 3.35 (2.71, 4.26) x1e1 | 3.85 (2.19, 7.41) x1e0 | -2.64 (-3.10, -2.22) x1e1 | -5.92 (-7.51, -4.71) x1e1 |
| Croatia | 1.74 (1.16, 2.53) x1e1 | 2.81 (2.01, 3.99) x1e0 | -7.98 (-10.38, -5.70) x1e0 | -1.70 (-2.20, -1.16) x1e1 |
| Lithuania | 1.18 (0.87, 1.50) x1e2 | 5.95 (4.39, 7.66) x1e0 | -6.81 (-8.85, -5.26) x1e0 | -1.59 (-2.47, -1.07) x1e1 |
| Uruguay | 2.34 (1.43, 3.66) x1e1 | 0 (0, 0) | -3.89 (-6.06, -2.33) x1e1 | -6.73 (-11.08, -3.91) x1e1 |
| Slovakia | 4.14 (3.05, 5.74) x1e1 | 1.65 (1.38, 1.94) x1e0 | -7.02 (-9.45, -5.58) x1e0 | -1.40 (-2.01, -1.07) x1e1 |
| Iceland | 1.78 (1.47, 2.23) x1e1 | 1.37 (1.06, 1.68) x1e0 | -8.26 (-10.89, -6.86) x1e0 | -1.73 (-2.32, -1.38) x1e1 |
| Guyana | 9.07 (7.23, 11.13) x1e0 | 0 (0, 0) | -2.06 (-2.62, -1.59) x1e1 | -3.90 (-4.94, -3.00) x1e1 |
| Puerto Rico | 1.29 (0.88, 1.77) x1e1 | 3.90 (2.72, 5.10) x1e0 | -7.89 (-9.64, -5.44) x1e0 | -1.72 (-2.23, -1.12) x1e1 |
| Seychelles | 8.34 (6.10, 11.30) x1e0 | 0 (0, 0) | -7.88 (-10.75, -5.44) x1e0 | -1.54 (-2.22, -1.02) x1e1 |
| Trinidad & Tobago | 2.39 (1.62, 3.39) x1e1 | 0 (0, 0) | -8.20 (-10.06, -6.49) x1e0 | -1.68 (-2.09, -1.29) x1e1 |
| Aruba | 6.62 (4.48, 10.25) x1e0 | 1.90 (1.30, 3.06) x1e0 | -4.50 (-8.03, -3.17) x1e0 | -1.02 (-1.84, -0.66) x1e1 |
| Antigua & Barbuda | 4.89 (3.68, 6.39) x1e0 | 0 (0, 0) | -1.64 (-2.00, -1.22) x1e0 | -3.22 (-4.10, -2.35) x1e0 |
| French Polynesia | 0 (0, 0) | 0 (0, 0) | -1.10 (-2.17, -0.43) x1e0 | -2.46 (-4.82, -0.91) x1e0 |
| Barbados | 0 (0, 0) | 0 (0, 0) | -1.01 (-1.43, -0.67) x1e0 | -2.28 (-3.27, -1.38) x1e0 |

**Table S6:** Percentage difference in number of TB cases among foreign-born individuals projected for 2025-2050 in each destination country under different scenarios, as compared to base-case. Countries ordered by total foreign-born TB notifications over 2014-2024. Positive values indicate additional TB cases as compared to the base-case. Negative values indicate fewer TB cases as compared to the base-case. Values in parentheses represent 95% uncertainty intervals.

| Country | Percentage difference in number of TB cases (%), by scenario |  |  |  |
| --- | --- | --- | --- | --- |
|  | USAID and Global Fund support ended in 2025 | USAID support ended in 2025 | Cautious optimism | Achieve End TB Strategy targets |
| United States | 101 (71, 132) | 5 (3, 6) | -29 (-37, -22) | -61 (-71, -50) |
| United Kingdom | 99 (72, 127) | 5 (3, 7) | -30 (-36, -23) | -61 (-70, -52) |
| Germany | 147 (113, 185) | 16 (12, 19) | -29 (-35, -23) | -60 (-69, -51) |
| France | 110 (85, 138) | 13 (10, 17) | -29 (-32, -24) | -65 (-71, -57) |
| Italy | 82 (57, 109) | 5 (4, 7) | -29 (-35, -22) | -58 (-67, -48) |
| South Korea | 46 (33, 62) | 7 (5, 10) | -30 (-37, -22) | -63 (-75, -50) |
| Spain | 69 (50, 92) | 8 (5, 13) | -29 (-34, -23) | -56 (-64, -47) |
| Japan | 40 (29, 50) | 5 (4, 7) | -30 (-36, -23) | -59 (-68, -49) |
| Saudi Arabia | 55 (37, 77) | 10 (6, 14) | -27 (-35, -19) | -59 (-71, -48) |
| Canada | 83 (58, 115) | 5 (3, 6) | -29 (-37, -21) | -63 (-74, -51) |
| Australia | 66 (46, 92) | 5 (3, 7) | -29 (-37, -21) | -61 (-72, -49) |
| Singapore | 38 (27, 60) | 9 (6, 11) | -31 (-36, -24) | -67 (-75, -57) |
| Qatar | 34 (21, 50) | 6 (3, 9) | -26 (-34, -17) | -54 (-68, -40) |
| Kuwait | 46 (30, 66) | 4 (3, 5) | -28 (-39, -18) | -60 (-78, -44) |
| Chile | 70 (45, 97) | 20 (12, 27) | -29 (-36, -21) | -58 (-70, -44) |
| Netherlands | 92 (71, 122) | 12 (9, 16) | -30 (-36, -24) | -65 (-74, -54) |
| Belgium | 218 (147, 318) | 19 (13, 27) | -30 (-35, -24) | -64 (-73, -53) |
| Sweden | 111 (87, 137) | 6 (5, 8) | -30 (-35, -23) | -62 (-70, -52) |
| Portugal | 38 (28, 48) | 2 (1, 3) | -30 (-36, -23) | -55 (-63, -46) |
| Switzerland | 100 (69, 154) | 7 (5, 10) | -30 (-35, -23) | -61 (-69, -52) |
| Austria | 100 (78, 123) | 7 (5, 9) | -29 (-34, -23) | -60 (-68, -50) |
| New Zealand | 44 (29, 62) | 4 (3, 6) | -29 (-37, -21) | -62 (-73, -51) |
| Hong Kong SAR China | 21 (14, 31) | 7 (5, 11) | -25 (-35, -17) | -58 (-71, -46) |
| Oman | 28 (17, 45) | 2 (1, 4) | -25 (-37, -16) | -56 (-71, -42) |
| Greece | 149 (108, 184) | 13 (10, 16) | -29 (-35, -23) | -61 (-70, -51) |
| Norway | 121 (91, 150) | 6 (4, 7) | -29 (-35, -23) | -61 (-69, -51) |
| Denmark | 99 (76, 124) | 6 (4, 7) | -30 (-35, -24) | -63 (-70, -54) |
| Israel | 149 (110, 194) | 12 (9, 15) | -30 (-37, -24) | -65 (-76, -54) |
| Poland | 123 (79, 171) | 9 (6, 13) | -27 (-34, -19) | -50 (-61, -40) |
| Czechia | 199 (124, 289) | 6 (4, 9) | -26 (-33, -19) | -48 (-59, -37) |

**Table S6 continued: Percentage difference in number of TB cases among foreign-born individuals projected for 2025-2050 in each destination country under different scenarios, as compared to base-case.**
| Country | Percentage difference in number of TB cases (%), by scenario |  |  |  |
| --- | --- | --- | --- | --- |
|  | USAID and Global Fund support ended in 2025 | USAID support ended in 2025 | Cautious optimism | Achieve End TB Strategy targets |
| Ireland | 132 (94, 167) | 3 (2, 4) | -30 (-36, -23) | -58 (-66, -48) |
| Bahrain | 37 (23, 56) | 3 (2, 4) | -27 (-36, -19) | -58 (-73, -45) |
| Finland | 143 (105, 184) | 7 (5, 10) | -29 (-35, -22) | -59 (-69, -48) |
| Malta | 63 (51, 77) | 5 (4, 7) | -27 (-31, -22) | -59 (-67, -49) |
| Brunei | 28 (19, 43) | 8 (5, 13) | -32 (-37, -26) | -70 (-78, -61) |
| Cyprus | 97 (69, 133) | 10 (7, 13) | -31 (-36, -24) | -65 (-73, -55) |
| Macao SAR China | 16 (10, 22) | 6 (4, 9) | -27 (-38, -17) | -60 (-77, -44) |
| Romania | 123 (69, 210) | 4 (2, 8) | -20 (-29, -12) | -41 (-56, -29) |
| Guam | 7 (5, 8) | 4 (3, 4) | -32 (-37, -26) | -70 (-83, -59) |
| Slovenia | 47 (36, 57) | 10 (8, 11) | -26 (-31, -21) | -59 (-77, -47) |
| Hungary | 137 (89, 190) | 6 (4, 9) | -26 (-35, -19) | -52 (-64, -40) |
| Luxembourg | 51 (27, 86) | 4 (2, 6) | -26 (-35, -18) | -51 (-64, -38) |
| Estonia | 424 (251, 638) | 24 (14, 35) | -23 (-33, -15) | -49 (-64, -36) |
| Latvia | 400 (246, 588) | 22 (14, 31) | -24 (-33, -16) | -50 (-63, -37) |
| Panama | 31 (21, 43) | 7 (5, 10) | -29 (-36, -21) | -61 (-70, -50) |
| Bahamas | 33 (27, 45) | 4 (2, 8) | -27 (-30, -24) | -61 (-68, -53) |
| Croatia | 54 (43, 71) | 9 (7, 12) | -26 (-29, -22) | -55 (-62, -46) |
| Lithuania | 392 (226, 610) | 20 (12, 30) | -22 (-32, -15) | -48 (-62, -36) |
| Uruguay | 20 (15, 24) | 0 (0, 0) | -33 (-40, -24) | -55 (-67, -43) |
| Slovakia | 160 (104, 223) | 6 (4, 9) | -27 (-35, -20) | -52 (-63, -41) |
| Iceland | 65 (49, 82) | 5 (4, 7) | -30 (-36, -24) | -62 (-72, -51) |
| Guyana | 11 (10, 12) | 1 (1, 2) | -26 (-29, -23) | -50 (-55, -44) |
| Puerto Rico | 51 (34, 77) | 15 (11, 22) | -31 (-38, -23) | -65 (-76, -52) |
| Seychelles | 30 (21, 39) | 1 (1, 1) | -28 (-37, -20) | -53 (-64, -41) |
| Trinidad & Tobago | 85 (59, 115) | 2 (1, 3) | -30 (-36, -24) | -60 (-70, -50) |
| Aruba | 46 (31, 67) | 13 (9, 18) | -31 (-39, -22) | -66 (-80, -53) |
| Antigua & Barbuda | 82 (61, 105) | 1 (1, 2) | -28 (-35, -21) | -55 (-64, -44) |
| French Polynesia | 15 (12, 19) | 6 (5, 8) | -32 (-39, -24) | -71 (-87, -54) |
| Barbados | 15 (11, 20) | 0 (0, 0) | -31 (-39, -23) | -66 (-78, -52) |

**Table S7:**
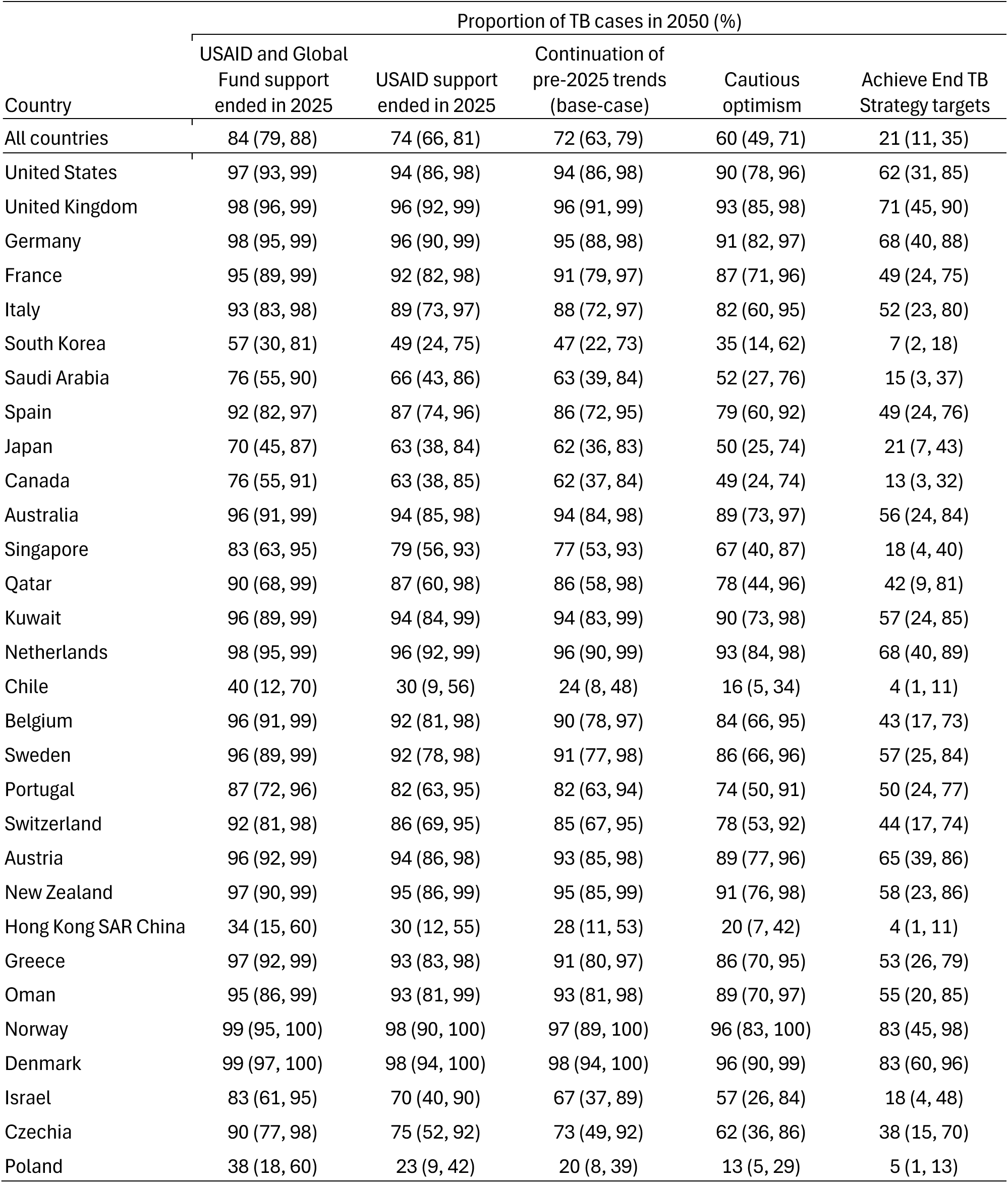
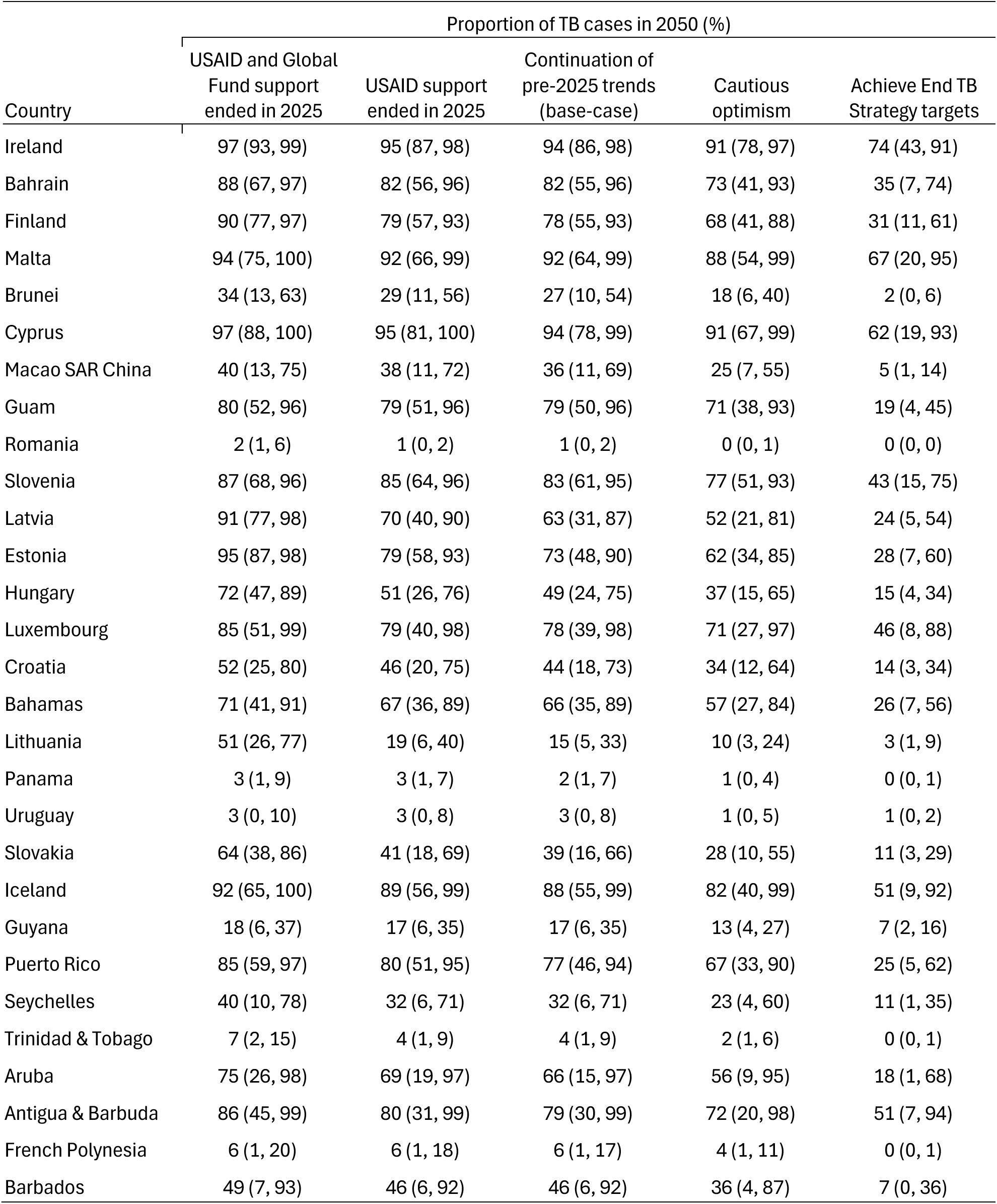
Projected proportion of TB cases in foreign-born population in 2050, by destination country and scenario. Countries ordered by total foreign-born TB notifications over 2014-2024. Values in parentheses represent 95% uncertainty intervals.

**Table S8:** Projected TB incidence rate in general population for 2050 in each destination country under different scenarios. Countries ordered by total foreign-born TB notifications over 2014-2024. Values in parentheses represent 95% uncertainty intervals.

| Country | TB incidence rate in 2050 (cases per 100,000 person-years) |  |  |  |  |
| --- | --- | --- | --- | --- | --- |
|  | USAID and Global Fund support ended in 2025 | USAID support ended in 2025 | Continuation of pre-2025 trends (base-case) | Cautious optimism | Achieve End TB Strategy targets |
| All countries | 11.5 (9.5, 14.0) | 7.1 (5.8, 8.9) | 6.5 (5.3, 8.3) | 4.6 (3.6, 6.1) | 2.3 (1.8, 3.0) |
| United States | 6.4 (4.2, 9.1) | 3.2 (2.1, 4.9) | 3.0 (1.9, 4.7) | 1.8 (1.1, 3.0) | 0.5 (0.2, 0.8) |
| United Kingdom | 20.3 (15.3, 25.5) | 10.1 (7.5, 14.0) | 9.5 (6.8, 13.3) | 5.7 (3.8, 8.8) | 1.4 (0.7, 2.5) |
| Germany | 14.3 (11.0, 18.9) | 6.9 (5.4, 8.9) | 5.7 (4.4, 7.5) | 3.5 (2.5, 4.9) | 0.9 (0.5, 1.6) |
| France | 25.1 (15.2, 37.2) | 14.7 (9.5, 23.4) | 12.6 (8.4, 22.0) | 8.5 (5.7, 14.8) | 2.1 (1.2, 3.8) |
| Italy | 11.5 (6.4, 22.1) | 6.9 (4.0, 16.5) | 6.5 (3.7, 15.0) | 4.1 (2.3, 8.1) | 1.4 (0.7, 2.6) |
| South Korea | 12.4 (7.2, 20.1) | 10.5 (5.9, 18.0) | 10.0 (5.6, 17.4) | 8.1 (4.2, 15.1) | 5.8 (2.3, 12.3) |
| Spain | 10.1 (6.5, 14.4) | 6.3 (4.3, 8.5) | 5.5 (3.9, 7.5) | 3.6 (2.5, 5.2) | 1.5 (0.8, 2.5) |
| Japan | 4.6 (2.8, 6.9) | 3.8 (2.3, 5.9) | 3.6 (2.2, 5.7) | 2.7 (1.5, 4.6) | 1.7 (0.8, 3.5) |
| Saudi Arabia | 8.0 (5.1, 12.6) | 5.7 (3.4, 9.2) | 5.3 (3.1, 8.1) | 4.0 (2.1, 6.5) | 2.3 (1.0, 4.5) |
| Canada | 14.4 (9.3, 20.4) | 9.4 (5.3, 14.8) | 9.1 (5.1, 14.3) | 6.7 (3.6, 11.7) | 4.0 (1.7, 8.3) |
| Australia | 11.0 (7.9, 15.2) | 6.7 (4.4, 9.7) | 6.3 (4.1, 9.2) | 3.8 (2.3, 6.0) | 0.9 (0.4, 1.6) |
| Singapore | 69.3 (42.0, 114.1) | 55.7 (31.0, 96.6) | 51.2 (28.6, 87.6) | 33.9 (18.8, 55.4) | 13.4 (5.6, 28.5) |
| Qatar | 22.0 (13.8, 39.1) | 16.5 (9.8, 29.9) | 15.3 (8.9, 27.9) | 9.9 (5.3, 17.6) | 3.5 (1.1, 9.6) |
| Kuwait | 27.2 (13.2, 54.8) | 17.9 (8.2, 39.6) | 17.1 (7.8, 39.1) | 10.3 (4.4, 22.4) | 2.2 (0.9, 4.3) |
| Chile | 16.2 (7.2, 30.1) | 13.6 (6.3, 26.2) | 12.6 (5.8, 25.4) | 11.4 (5.0, 24.5) | 10.2 (3.8, 23.4) |
| Netherlands | 12.0 (9.0, 18.4) | 6.7 (4.6, 10.6) | 5.8 (3.9, 9.1) | 3.6 (2.3, 5.7) | 0.7 (0.4, 1.3) |
| Belgium | 30.0 (19.0, 46.5) | 12.4 (8.5, 17.6) | 10.0 (7.0, 13.7) | 6.3 (4.4, 9.2) | 1.7 (0.8, 3.2) |
| Sweden | 15.1 (10.4, 25.8) | 7.8 (5.1, 13.4) | 7.2 (4.7, 12.8) | 4.6 (2.9, 7.8) | 1.4 (0.7, 2.5) |
| Portugal | 15.6 (10.1, 27.0) | 11.8 (7.2, 21.0) | 11.6 (7.0, 20.9) | 7.7 (4.4, 13.2) | 3.9 (2.0, 7.0) |
| Switzerland | 15.5 (11.6, 20.2) | 8.6 (6.2, 11.8) | 8.0 (5.7, 11.0) | 5.3 (3.6, 7.9) | 2.1 (1.1, 4.0) |
| Austria | 11.0 (8.3, 13.9) | 6.2 (4.8, 8.3) | 5.7 (4.3, 7.8) | 3.6 (2.5, 5.1) | 1.1 (0.6, 1.9) |
| New Zealand | 10.1 (7.2, 14.7) | 7.1 (4.6, 11.1) | 6.7 (4.2, 10.6) | 4.0 (2.2, 6.3) | 0.8 (0.3, 1.6) |
| Hong Kong SAR China | 25.2 (13.0, 46.9) | 23.9 (11.8, 45.4) | 23.2 (11.2, 44.6) | 21.0 (9.2, 41.9) | 17.8 (6.5, 39.0) |
| Oman | 5.6 (2.3, 10.9) | 4.0 (1.6, 8.2) | 3.9 (1.6, 8.1) | 2.3 (1.0, 4.6) | 0.5 (0.2, 1.1) |
| Greece | 9.5 (6.3, 16.9) | 4.2 (3.0, 6.3) | 3.6 (2.4, 5.3) | 2.2 (1.4, 3.3) | 0.6 (0.3, 1.1) |
| Norway | 15.0 (9.5, 22.0) | 6.6 (4.3, 9.8) | 6.2 (4.0, 9.2) | 3.7 (2.3, 5.6) | 0.9 (0.4, 1.6) |
| Denmark | 12.5 (9.1, 16.6) | 6.6 (4.8, 8.6) | 6.1 (4.5, 8.0) | 3.7 (2.6, 5.3) | 0.8 (0.4, 1.4) |
| Israel | 8.7 (4.8, 14.6) | 4.6 (2.6, 8.0) | 4.2 (2.3, 7.5) | 3.2 (1.6, 5.9) | 1.7 (0.6, 4.2) |
| Poland | 3.3 (1.8, 6.0) | 2.7 (1.3, 5.4) | 2.6 (1.2, 5.3) | 2.4 (1.0, 5.2) | 2.2 (0.8, 5.0) |
| Czechia | 7.4 (3.4, 19.7) | 2.3 (1.3, 4.1) | 2.1 (1.2, 4.0) | 1.5 (0.8, 2.5) | 0.9 (0.4, 1.6) |

**Table S8 continued: Projected TB incidence rate in general population for 2050 in each destination country under different scenarios.**
| Country | TB incidence rate in 2050 (cases per 100,000 person-years) |  |  |  |  |
| --- | --- | --- | --- | --- | --- |
|  | USAID and Global Fund support ended in 2025 | USAID support ended in 2025 | Continuation of pre-2025 trends (base-case) | Cautious optimism | Achieve End TB Strategy targets |
| Ireland | 10.1 (5.6, 16.5) | 4.7 (3.0, 7.0) | 4.5 (2.8, 6.8) | 2.8 (1.7, 4.4) | 1.0 (0.4, 1.7) |
| Bahrain | 14.0 (8.1, 21.5) | 9.7 (5.3, 16.4) | 9.4 (5.0, 16.0) | 6.4 (3.2, 11.8) | 2.5 (0.9, 6.1) |
| Finland | 7.5 (4.0, 11.7) | 3.5 (2.0, 5.3) | 3.2 (1.9, 4.7) | 2.2 (1.3, 3.4) | 1.0 (0.5, 2.0) |
| Malta | 14.3 (8.9, 21.5) | 10.3 (6.6, 17.0) | 9.7 (6.1, 15.9) | 6.6 (4.0, 11.0) | 2.1 (0.9, 6.0) |
| Brunei | 125.6 (59.2, 244.9) | 118.0 (54.6, 237.7) | 115.1 (52.3, 235.4) | 103.0 (43.0, 222.6) | 88.2 (29.0, 206.8) |
| Cyprus | 10.1 (7.0, 13.4) | 6.0 (4.0, 8.4) | 5.3 (3.5, 7.6) | 3.2 (2.0, 4.9) | 0.7 (0.3, 1.6) |
| Macao SAR China | 41.3 (18.6, 78.5) | 39.2 (17.8, 75.7) | 37.9 (17.0, 73.8) | 32.2 (13.4, 67.0) | 25.7 (9.1, 60.6) |
| Romania | 12.1 (4.6, 27.7) | 11.9 (4.4, 27.6) | 11.9 (4.4, 27.6) | 11.9 (4.4, 27.6) | 11.8 (4.3, 27.5) |
| Guam | 72.5 (34.0, 155.0) | 71.2 (33.1, 153.9) | 68.8 (32.1, 148.6) | 46.9 (22.0, 96.4) | 16.3 (4.8, 46.5) |
| Slovenia | 2.5 (1.4, 3.9) | 2.1 (1.2, 3.4) | 1.9 (1.1, 3.2) | 1.4 (0.8, 2.3) | 0.5 (0.2, 1.1) |
| Hungary | 1.6 (1.0, 2.4) | 0.9 (0.5, 1.6) | 0.9 (0.5, 1.5) | 0.7 (0.4, 1.3) | 0.5 (0.2, 1.2) |
| Luxembourg | 3.8 (2.1, 6.7) | 2.7 (1.4, 5.3) | 2.6 (1.3, 5.2) | 1.8 (0.8, 4.3) | 1.0 (0.3, 3.3) |
| Estonia | 14.4 (8.2, 20.0) | 3.5 (2.2, 5.1) | 2.6 (1.6, 4.1) | 1.9 (1.0, 3.2) | 1.0 (0.4, 2.1) |
| Latvia | 13.6 (8.6, 20.4) | 3.8 (2.3, 6.3) | 3.1 (1.7, 5.5) | 2.3 (1.1, 4.8) | 1.5 (0.5, 3.8) |
| Panama | 35.5 (10.2, 90.1) | 35.3 (10.1, 89.9) | 35.2 (10.0, 89.8) | 35.0 (9.8, 89.6) | 34.7 (9.5, 89.3) |
| Bahamas | 13.1 (7.3, 21.9) | 11.3 (6.2, 20.3) | 10.8 (6.0, 19.4) | 8.6 (4.7, 16.9) | 5.1 (1.9, 13.2) |
| Croatia | 1.3 (0.6, 2.3) | 1.1 (0.6, 2.1) | 1.1 (0.5, 2.0) | 0.9 (0.4, 1.8) | 0.7 (0.3, 1.6) |
| Lithuania | 7.2 (4.4, 12.6) | 4.4 (1.9, 9.8) | 4.2 (1.7, 9.6) | 4.0 (1.5, 9.4) | 3.8 (1.3, 9.3) |
| Uruguay | 94.1 (33.6, 214.0) | 93.7 (33.5, 213.8) | 93.7 (33.5, 213.8) | 92.8 (33.2, 213.6) | 92.1 (32.2, 213.2) |
| Slovakia | 1.1 (0.7, 1.7) | 0.7 (0.3, 1.2) | 0.6 (0.3, 1.2) | 0.5 (0.2, 1.1) | 0.5 (0.2, 1.0) |
| Iceland | 6.4 (4.1, 11.4) | 4.6 (2.7, 9.2) | 4.3 (2.5, 8.9) | 2.7 (1.5, 5.8) | 0.9 (0.3, 3.6) |
| Guyana | 18.8 (8.4, 39.2) | 18.5 (8.3, 39.0) | 18.5 (8.2, 38.9) | 17.7 (7.4, 38.1) | 16.8 (6.4, 37.0) |
| Puerto Rico | 0.5 (0.3, 0.8) | 0.4 (0.2, 0.6) | 0.3 (0.2, 0.5) | 0.2 (0.1, 0.4) | 0.1 (0.0, 0.3) |
| Seychelles | 31.4 (9.9, 87.8) | 28.4 (7.6, 84.5) | 28.3 (7.5, 84.4) | 25.6 (5.9, 83.0) | 23.1 (4.0, 79.8) |
| Trinidad & Tobago | 21.6 (8.0, 47.3) | 21.1 (7.5, 46.9) | 21.1 (7.5, 46.9) | 20.8 (7.1, 46.6) | 20.5 (6.8, 46.3) |
| Aruba | 11.1 (5.0, 27.7) | 8.8 (3.6, 24.5) | 8.0 (3.0, 23.6) | 6.1 (1.8, 22.5) | 4.0 (0.3, 20.3) |
| Antigua & Barbuda | 4.2 (2.1, 10.5) | 2.6 (1.1, 6.3) | 2.6 (1.1, 6.2) | 1.8 (0.7, 5.0) | 1.0 (0.2, 4.0) |
| French Polynesia | 15.1 (4.2, 39.4) | 15.1 (4.2, 39.3) | 15.0 (4.2, 39.3) | 14.8 (3.9, 39.1) | 14.5 (3.5, 38.8) |
| Barbados | 1.6 (0.4, 6.8) | 1.5 (0.3, 6.7) | 1.5 (0.3, 6.7) | 1.3 (0.2, 6.5) | 1.1 (0.1, 6.0) |

